# Assessing the effects of non-pharmaceutical interventions on SARS-CoV-2 transmission in Belgium by means of an extended SEIQRD model and public mobility data

**DOI:** 10.1101/2020.07.17.20156034

**Authors:** Tijs W. Alleman, Jenna Vergeynst, Lander De Visscher, Michiel Rollier, Elena Torfs, Ingmar Nopens, Jan M. Baetens

**Affiliations:** BIOMATH, Department of Data Analysis and Mathematical Modelling, Ghent University, Coupure links 653, 9000 Gent, Belgium; KERMIT, Department of Data Analysis and Mathematical Modelling, Ghent University, Coupure links 653, 9000 Gent, Belgium; Department of Epidemiology and Public Health, Sciensano, BE-1050 Brussels, Belgium

**Keywords:** SARS-CoV-2, age-stratified compartmental SEIQRD model, non-pharmaceutical interventions, Google Community Mobility data, effective reproduction number, model calibration, schools closure

## Abstract

We present a compartmental extended SEIQRD metapopulation model for SARS-CoV-2 spread in Belgium. We demonstrate the robustness of the calibration procedure by calibrating the model using incrementally larger datasets and dissect the model results by computing the effective reproduction number at home, in workplaces, in schools, and during leisure activities. We find that schools are an important transmission pathway for SARS-CoV-2, with the potential to increase the effective reproduction number from *R*_*e*_ = 0.66 ± 0.04 (95 % CI) to *R*_*e*_ = 1.09 ± 0.05 (95 % CI) under lockdown measures. The model accounts for the main characteristics of SARS-CoV-2 transmission and COVID-19 disease and features a detailed representation of hospitals with parameters derived from a dataset consisting of 22 136 hospitalized patients. Social contact during the pandemic is modeled by scaling pre-pandemic contact matrices with Google Community Mobility data and with effectivity-of-contact parameters inferred from hospitalization data. The calibrated social contact model with its publically available mobility data, although coarse-grained, is a readily available alternative to social-epidemiological contact studies under lockdown measures, which were not available at the start of the pandemic.

## 1 Introduction

After an initial outbreak in early 2020 in Wuhan, China, *Severe acute respiratory syndrome coronavirus 2* (SARS-CoV-2) has spread globally [1]. SARS-CoV-2 is capable of sustained human-to-human transmission [2] and may cause severe disease and death, especially in older individuals. The SARS-CoV-2 pandemic has, in general, shown a remarkably low incidence among children and young adults [3, 4, 5]. Furthermore, presymptomatic transmission is a major contributor to SARS-CoV-2 spread [6, 7]. Both on March 15th, 2020, and on October 19th, 2020, the Belgian governments imposed social restrictions after testing & tracing methods had failed to prevent the large-scale spread of SARS-CoV-2. Recently, pharmaceutical interventions under the form of vaccinations have become available. If natural immunity wanes or if SARS-CoV-2 further mutates, it is expected that SARS-CoV-2 will become endemic [8]. Hence, there is a need for well-informed models and knowledge build-up to assist policymakers in choosing the best non-pharmaceutical and pharmaceutical interventions during future SARS-CoV-2 outbreaks.

Currently, four other models exist to inform policymakers in Belgium. The agent-based model (ABM) of Willem et al. [9], the data-driven model by Barbe et al. [10] and the nation-level, age-stratified compartmental models of Abrams et al. [11] and Franco [12]. The models of Abrams et al. [11] and Franco [12] feature similar disease dynamics as our model but rely on different assumptions to model social contact. The different model outputs are currently combined into an ensemble to inform policymakers [13]. In the ensemble, each model fulfills a niche, for instance, the ABM of Willem et al. [9] is good for studying microscopic social behavior, and was used to inform the optimal *household bubble* size. The model of Barbé excels at short-term forecasts while our model, together with the compartmental models of Abrams et al. [11] and Franco [12], are well-fit to study the long-term effects of population-wide interventions.

In this work, we built a compartmental, age-stratified, nation-level model which accounts for the main characteristics of SARS-CoV-2 disease. The model features a detailed representation of hospitals with residence times and mortalities derived from a large dataset of hospitalized patients in Belgium. We built a social contact model which scales pre-pandemic contact matrices from a study by Willem et al. [14] with the Google Community Mobility data [15] and with effectivity-of-contact parameters derived from hospitalization data using an *Markov Chain Monte Carlo* (MCMC) method [16]. Tardiness in compliance with social restrictions is included using a delayed-ramp model and waning of humoral immunity is included by estimating the rate of seroreversion from two serological datasets. We find that the combination of the deterministic epidemiological model, which incorporates rigid a priori knowledge on disease dynamics, and the calibrated effectivity-of-contact parameters in the social contact model allows us to combine the ease of long-term extrapolation and scenario analysis of compartmental models with the flexibility of a data-driven model. The model does not require ad hoc tweaking and is computationally cheap, making it ideal to perform optimizations that require thousands of model evaluations. Further, due to the public nature of the Google Community Mobility data, the model provides a more rapidly deployable alternative to social epidemiological studies comparing mixing patterns during and after lockdown, such as Coletti et al. [17] for Belgium, which were not available at the start of the pandemic.

Using a hospitalization dataset of 22 136 *coronavirus disease 19* (COVID-19) patients in Belgian hospitals, we computed age-stratified hospital residence times and mortalities. Using the obtained parameters, we found the model was able to predict the total number of patients and the number of deceased patients in Belgian hospitals well. We calibrated the model to hospitalization data made publically available by the Belgian *Scientific Institute of Public Health* (Sciensano) and demonstrated the calibration procedure’s robustness. We computed the basic reproduction numbers (*R*_0_) and the time to reach compliance to lockdown measures during both *coronavirus disease 2019* (COVID-19) waves in Belgium. The average time to for anti-SARS-CoV-2 antibodies to wane (seroreversion), was estimated as 9.2 months (IQR: 7.2 - 12.1 months). Using the calibrated model, we computed the relative share of contacts and the effective reproduction numbers and found these to be in line with estimates from other authors at home, at school, at work and during leisure activities to asses their effect on SARS-CoV-2 spread during both 2020 COVID-19 waves. We observed a strong correlation between school re-opening and increases in SARS-CoV-2 transmission. More precisely, schools have the potential to increase the effectiveic reproduction number from *R*_*e*_ = 0.67 ± 0.04 (95 % CI) to *R*_*e*_ = 1.09 ± 0.05 (95 % CI) under lockdown measures.

Throughout the work, Belgium is used as a case but the scope of the work is extendable to other countries. Since February 2021, the effects of new SARS-CoV-2 strains and pharmaceutical interventions (vaccines) need to be accounted for. For this purpose, a model extension was developed and is currently used in the aforementioned model ensemble [13]. However, due to the longevity of this work, we chose to limit the scope of this study to the effects of non-pharmaceutical interventions.

## 2 Materials and methods

### 2.1 The extended SEIQRD-model

#### 2.1.1 Disease dynamics

The SEIR(D) model [18] is a compartmental model that subdivides the human population into four groups: 1. susceptible individuals (S), 2. exposed individuals in the latent phase (E), 3. infectious individuals capable of transmitting the disease (I) and 4. individuals removed from the population either through immunization or death (R/D). Despite being a simple and idealized reality, the SEIR(D) dynamics are used extensively to predict the outbreak of infectious diseases and this was no different during the SARS-CoV-2 outbreak earlier this year [1, 3, 19].

In this work, we extended the SEIRD model to incorporate more expert knowledge on SARS-CoV-2 disease dynamics. For that purpose, the infectious compartment was split into four parts. The first is a period of presymptomatic infectiousness because several studies have shown that presymptomatic transmission is a dominant transmission mechanism of SARS-CoV-2 [6, 7]. After the presymptomatic period, three possible infectious outcomes are modeled: (1) Asymptomatic outcome, for individuals who show no symptoms at all, (2) Mild outcome, for individuals with mild symptoms who recover at home, and (3) Hospitalization, when mild symptoms worsen. Children and young adults have a high propensity to experience an asymptomatic or mild outcome, while older individuals have a higher propensity to be hospitalized [6, 7]. Belgian hospitals generally have two wards for COVID-19 patients: 1) *cohort*, where patients are not monitored continuously and 2) *Intensive care units* (ICUs), for patients with the most severe symptoms. Intensive care includes permanent monitoring, the use of ventilators, or the use of extracorporeal membrane oxygenation (ECMO). Patients can perish in both hospital wards, but mortalities are generally lower in cohort. After a stay in an ICU, patients return to cohort for recovery in the hospital. During the recovery stay, mortality is limited. We assume that mildly infected individuals and hospitalized patients cannot infect susceptibles are thus quarantined. Because reinfections with SARS-CoV-2 have already been reported [20, 21, 22, 23], and because it has already been estimated that anti-SARS-CoV-2 antibodies wane [24, 25], we incorporate waning antibody immunity by sending recovered individuals back to the susceptible population pool. The model dynamics are depicted in Figure 1.

**Figure 1:**
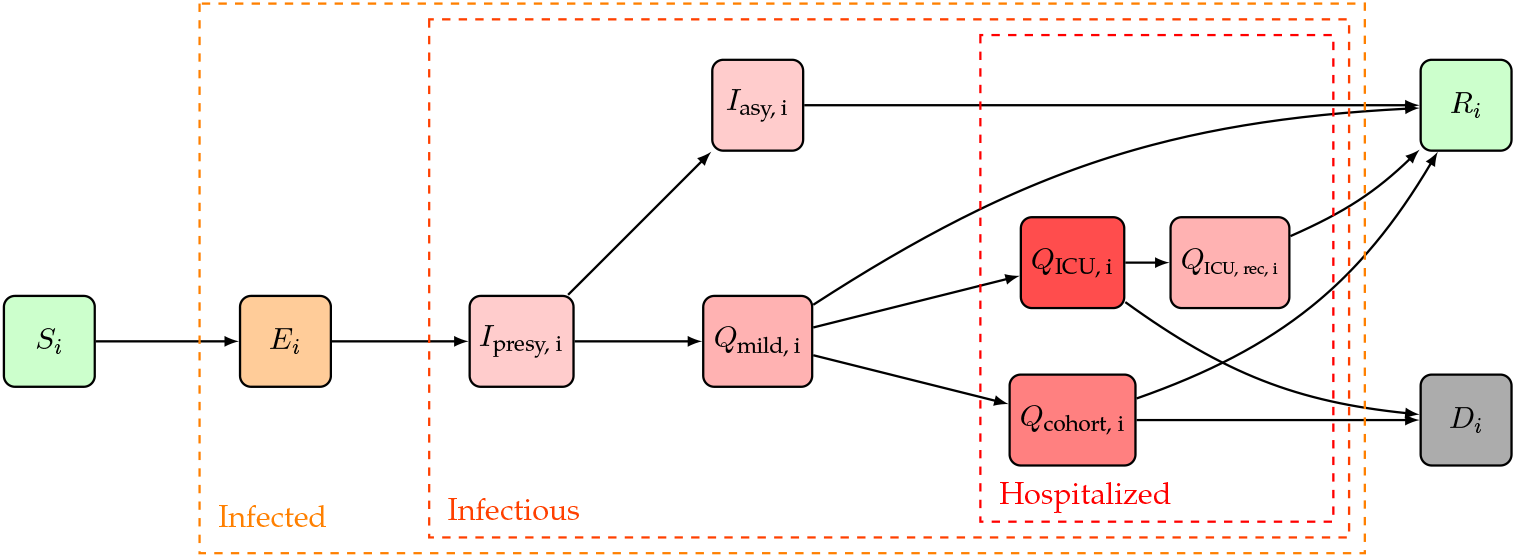
Extended SEIQRD dynamics used in this study. Here, *S* stands for susceptible, *E* for exposed, *I*_presy_ for presymptomatic and infectious, *I*_asy_ for asymptomatic and infectious, *Q*_mild_ for mildly symptomatic and infectious, *Q*_cohort_ for cohort, *Q*_ICU,rec_ for a recovery stay in cohort coming from IC, *Q*_ICU_ for Intensive Care Unit, *D* for dead and *R* for recovered. A subscript *i* is used to denote the *i*th age strate of the model, the model has a total of nine age strata. An overview of the model parameters can be found in table 1.

#### 2.1.2 Model structure and equations

In this work, we implemented the extended SEIQRD dynamics shown in Figure 1 using ordinary differential equations (ODEs), without spatial stratification and with age-stratification.

This was accomplished by defining a system of *K* × *N* ordinary differential equations, one for every of the *K* = 10 model compartments, each of which is further split into *N* = 9 age-stratified metapopulations. The age groups have different contact rates with other age groups and the disease progresses differently for each age group, making the model behave realistically. Our model consists of 9 age classes, i.e., [0, 10(, [10, 20(, [20, 30(, [30, 40(, [40, 50(, [50, 60(, [60, 70(, [70, 80(, [80, ∞(. The advantage of using ODEs over network- or agent-based models are the limited computational resources required to explore scenarios and perform optimizations that require thousands of function evaluations. The disadvantage is the assumption of homogeneous mixing, i.e. every individual is equally likely to come into contact with another individual. More realistic approaches are spatial patch models, network-based models, agent-based models, or combinations thereof. However, these come at a substantial computational cost. Because Belgium is a small and heavily urbanized country, a spatially explicit model becomes relevant at very low SARS-CoV-2 prevalence. The macroscopic coarse-graining of homogeneous mixing works well to describe major COVID-19 waves but is less fit for monitoring the disease at low prevalence. The model dynamics are translated into the following system of coupled ordinary differential equations,

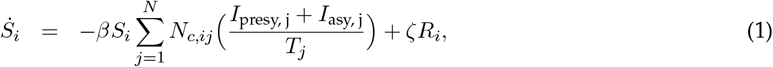

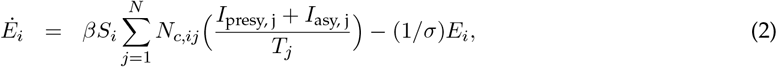

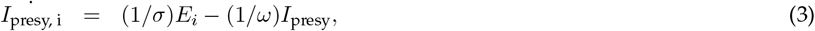

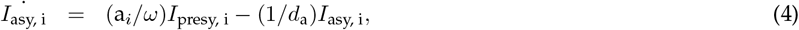

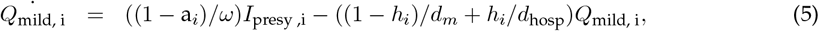

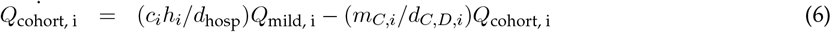

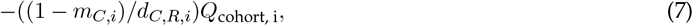

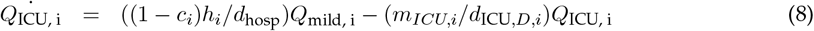

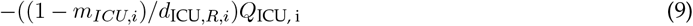

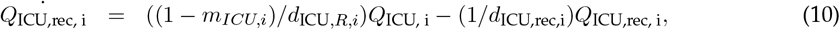

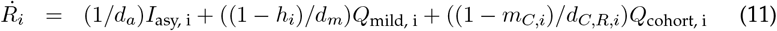

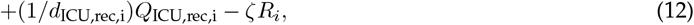

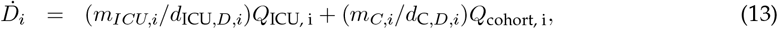

for *i* = 1, 2, …, 9. Here, *T* stands for total population (Table 1), *S* stands for susceptible, *E* for exposed, *I*_presy_ for presymptomatic and infectious, *I*_asy_ for asymptomatic and infectious, *Q*_mild_ for mildly symptomatic and infectious, *H*_cohort_ for cohort, *H*_ICU,rec_ for a recovery stay in cohort coming from Intensive Care, *H*_ICU_ for Intensive Care Unit, *D* for dead and *R* for recovered. A subscript to these variables is used to refer to one of the nine age strata in the model. Using the above notation, all model states are 9-dimensional vectors,

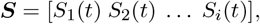

where *S*_*i*_(*t*) denotes the number of susceptibles in age-class *i* at time *t* after the introduction of SARS-CoV-2 in the population. As initial condition, the whole population is assumed susceptible to SARS-CoV-2 and one exposed individual and one pre-symptomatic infectious individual in every age class is assumed, so *E*_*i*_(0) = *I*_*i*_(0) = 1 for all *i* = 1, 2, …, 9. The time between the start of the simulation and the start of data collection is then estimated when calibrating the model. An overview of all model parameters, their values, and their meaning can be found in table 1. In what follows, the most important model parameters and their chosen values are motivated.

**Table 1:** Overview of simulation parameters used in the extended SEIQRD metapopulation model.

| Symbol | Parameter | Value | Unit | Reference |
| --- | --- | --- | --- | --- |
| $a$ | subclinical fraction per age group | [0.98 0.98 0.88 0.69 0.59 0.39 0.13 0.07 0.01]<br>population mean: 0.57 | (—) | Wu et al. [37] |
| $h$ | fraction of mildly infected individuals requiring hospitalisation | [0.01 0.02 0.02 0.02 0.02 0.05 0.11 0.22 0.57]<br>population mean: 0.08 | (—) | Inferred |
| $c$ | fraction of hospitalisations not requiring ICU transfer | Table 4, population mean: 0.84 | (—) | Hospital dataset |
| $d_a$ | duration of subclinical infection | 6.54 | days | Inferred |
| $d_m$ | duration of mild infection | 7 | days | To et al. [32] |
| $d_{\text{hosp}}$ | average time from symptom onset to hospitalization | Table 6, population mean: 6.4 | days | Hospital dataset |
| $d_{C,R}$ | length of cohort stay if recovered | Table 5, population mean: 10.8 | days | Hospital dataset |
| $d_{C,D}$ | length of cohort stay if deceased | Table 5, population mean: 11.8 | days | Hospital dataset |
| $d_{\text{ICU},R}$ | length of ICU stay if recovered | Table 5, population mean: 12.0 | days | Hospital dataset |
| $d_{\text{ICU},D}$ | length of ICU stay if deceased | Table 5, population mean: 15.2 | days | Hospital dataset |
| $d_{\text{ICU},\text{rec}}$ | length of recovery and observation stay in cohort after ICU stay | Table 6, population mean: 11.2 | days | Hospital dataset |
| $m_C$ | mortality in cohort | Table 4, population mean: 0.17 | (—) | Hospital dataset |
| $m_{\text{ICU}}$ | mortality in ICU | Table 4, population mean: 0.46 | (—) | Hospital dataset |
| $\sigma$ | length of latent period | 4.5 | days | Computed |
| $\omega$ | length of presymptomatic infectious period | 0.7 | days | Wei et al. [7], He et al. [29] |
| $\sigma + \omega$ | length of incubation period | 5.2 | days | Liu et al. [6] |
| $\beta$ | probability of infection upon contact with an individual capable of transmitting SARS-CoV-2 under the assumption that the infectee is 100 % susceptible to SARS-CoV-2 infection | 0.032 | (—) | Inferred |
| $T_0$ | total population | [1.31 1.30 1.40 1.50 1.52 1.60 1.35 0.91 0.66]*1e6,<br>total population: 11.54 * 1e6 | people | StatBEL [54] |
| $N_c$ | contact matrix | 9x9 matrix | days<sup>-1</sup> | Willem et al. [14] |

#### 2.1.3 Model parameters

##### Transmission rate and social contact data

The transmission rate of the disease depends on the product of four contributions (Equation 1). The first contribution, (*I*_presy, j_ + *I*_asy, j_)*/T*_*j*_, is the fraction of contagious individuals in age group *j*. We thus assume presymptomatic and asymptomatic individuals spread the disease, while mildly infected are assumed to self-quarantine and hospitalized individuals cannot infect health care workers. The second contribution, *N*_*c,ij*_, is the average number of human-to-human interactions of an individual in age group *i*, with an individual in age group *j* per day. The sum of the first two contributions over all age groups 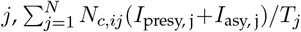, is the number of contacts of an individual in age group *i* that can result in SARS-CoV-2 transmission. This is multiplied with the number of susceptibles in age group *i* (*S*_*i*_), and with *β*, the probability of contracting COVID-19 when encountering a contagious individual, to compute the number of effective contacts at every timestep. We assume that the per-contact transmission probability *β* is independent of age and we infer its value by calibrating our model to Belgian hospitalization data. In a model-based inference-based study by Davies et al. [3], it was deduced that children were less susceptible to SARS-CoV-2 disease. Viner et al. [26] analyzed 32 studies that reported on the susceptibility of children and found preliminary evidence that susceptibility to SARS-CoV-2 infection is lower in children. However, it assumed in our model that individuals of all ages to have an equal susceptibility to and transmissibility of SARS-CoV-2. The number of (pre-pandemic) human-human interactions, ***N***_*c*_, are both place and age-dependent. These matrices assume the form of a 9×9 *interaction matrix* where an entry *i, j* denotes the number of social contacts age group *i* has with age group *j* per day. These matrices are available for homes (***N***_c, home_), schools (***N***_c, schools_), workplaces (***N***_c, work_), in public transport (***N***_c, transport_), during leisure activities (***N***_c, leisure_) and during other activities (***N***_c, others_), from a study by Willem et al. [14]. The total number of prepandemic social interactions must be translated into an appropriately weighted sum of the contributions in different places, adequately describing pandemic social behavior (Section 2.3). The basic reproduction number *R*_0_, defined as the expected number of secondary cases directly generated by one case in a population where all individuals are susceptible to infection, is computed using the next-generation matrix (NGM) approach introduced by Diekmann et al. [27, 28]. For our model, the basic reproduction number of age group *i* is,

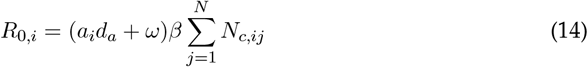

and the population basic reproduction number is calculated as the weighted average over all age groups using the demographic data in Table 1. The detailed algebra underlying the computation of Equation 14 is presented in the supplementary materials (Section A.4).

##### Infectiousness

The duration of infectiousness is determined by the number of days patients can spread viral particles. Several studies have reported patients have the highest viral load of the coronavirus at the time they are diagnosed and patient’s viral loads declining gradually over time [29, 30, 31, 32, 33]. He et al. [29] inferred the infectiousness profile of COVID-19 patients to be an approximately normal distribution, with the peak infectivity roughly at the time of symptom onset and infectiousness quickly declining within 7 days after symptom onset. A comparison of viral load between symptomatic and one asymptomatic case revealed similar viral loads, an indicator that asymptomatic individuals can be as infectious as symptomatic patients [33]. He et al. [29] further concluded that 44 % of secondary cases were infected during the presymptomatic stage, a finding consistent with studies from other authors [6, 7]. Wei et al. [7] determined that presymptomatic transmission exposure occurred 1-3 days before the source patient developed symptoms. In Equation 2, *s* denotes the length of the latent, non-infectious period and in Equation 3, *ω* is the length of the presymptomatic infectious period. In this work, we assume the incubation period, equal to *ω* + *s*, lasts 5.2 days [6]. The length of the presymptomatic period is fixed at 0.7 days, which corresponds to 44 % of SARS-CoV-2 infections experiencing a presymptomatic infectious period of 2 days. The duration of infectiousness for mildly symptomatic cases (*d*_*m*_) is assumed to be 7 days. The average duration of asymptomatic infectiousness, on which the basic reproduction number (*R*_0_) depends (Equation 14), will be inferred from hospitalization data using an MCMC method (Section 2.4).

##### Disease severity and hospitalizations

The model parameter *a*_*i*_ (Equation 4) is the probability of an individual in age group *i* having a subclinical infection. Several authors have attempted to estimate the fraction of asymptomatic infections. Li et al. [34] estimated that 86 % of coronavirus infections in China were *undocumented* in the weeks before their government instituted stringent quarantines. However, this figure includes an unknown number of mildly symptomatic cases and is thus an overestimation of the asymptomatic fraction. In Iceland, citizens were invited for testing regardless of symptoms. Of all people with positive test results, 43 % were subclinical [35]. A systematic review and meta-analysis by Buitrago-Garcia et al. [36] suggested a lower subclinical fraction of 31 % (26 % - 37 %, 95 % CI). If the subclinical fractions per age group estimated by Davies et al. [3] are applied to the Belgian population, an average subclinical fraction of 57 % is obtained for Belgium. In this study, we applied the relative subclinical fraction per age group of Wu et al. [37] to obtain a population average subclinical fraction of 57 % (Table 1). In Equation 5, *h* is the fraction of mild cases that require hospitalization and in Equation 8, *c* is the fraction of the hospitalized which remain in cohort. In this study, the age-stratified hospitalization probabilities (*h*) were inferred from hospital mortality data (Table 1) and the age-stratified distributions between cohort and ICU (*c*) were computed using data from 22 136 patients treated in Belgian hospitals (Section 2.2). In Equation 5, *d*_hosp_ is the average time between first symptoms and hospitalization, which was previously estimated as 5-9 days by Linton et al. [38] and as 4 days by To et al. [32]. In Equations 8, 9 and 10, *d*_C,R_, *d*_C,D_, *d*_ICU,R_ and *d*_ICU,D_ are the age-stratified average lengths of a hospital stay in cohort and in an ICU. The subscript *R* denotes the duration if the patient recovers, while subscript *D* denotes the duration if the patient perishes. *m*_*C*_ and *m*_ICU_ are the age-stratified mortalities of patients in cohort and in ICU. In Equation 10, *d*_ICU,rec_ denoted the age-stratified length of a recovery stay in cohort after a stay in ICU. The aforementioned hospitalization parameters are computed using data from 22 136 patients treated in Belgian hospitals. The methodology of the analysis is presented in Section 2.2, the results of the analysis are presented in Section 3.1

##### Testing, tracing and quarantine, waning antibody immunity

The effects of testing, tracing and quarantine are not explicitly implemented for this study. Reinfections with SARS-CoV-2 have been reported in single cases in the USA [20], Ecuador [21] and Belgium [22]. Further, two asymptomatic reinfections were also reported in Indian healthcare workers [23]. Rosado et al. [24] estimated that antibodies could wane in 50% of recovered individuals after 1 year. Wheatley et al. [25] found that both neutralizing and binding antibody responses decay after recovery from a mild COVID-19 infection. Although the long-term kinetics of the antibody response to SARS-CoV-2 will not be definitively quantified until infected individuals are followed years after a confirmed infection, and although the persistence of serum antibodies is unlikely to be the sole determinant of long-lasting immunity (memory T and B cells), it is clear that waning of antibodies best be included in our model. In Equations 1 and 12, the rate of anti-SARS-CoV-2 antibody waning is denoted as *ζ*, and its inverse is the average time for anti-SARS-CoV-2 antibodies to wane. Using serological data by Herzog et al. [39] and the *Belgian Scientific Institute of Public Health* (Sciensano), the distribution of *ζ* will be inferred using an MCMC method.

### 2.2 Analysis of hospital surveillance data

A subset of data from the Belgian COVID-19 clinical surveillance on hospitalizations by Van Goethem et al. [40], which was anonymized and provided through a secured data transfer platform by the *Belgian Scientific Institute of Public Health* (Sciensano), is analyzed to compute age-stratified estimates of the following model parameters: the distribution between the cohort and IC wards (*c*), the residence times in the cohort and IC wards, in the case of recovery and in the case of death (*d*_*C,R*_, *d*_*C,D*_, *d*_ICU,*R*_, *d*_ICU,*D*_), the residence time for a recovery stay in cohort after a stay in ICU (*d*_ICU,rec_), the time between symptom onset and hospitalization (*d*_hospital_) and the mortalities in the hospital, cohort and IC wards (*m*_C,ICU_, *m*_C_, *m*_ICU_). The raw data consistes of 52 327 patients hospitalized in Belgian hospitals between March 4th, 2020, and March 3rd, 2021. Data are reported for all hospitalized patients with a confirmed COVID-19 infection (diagnosed using reverse transcriptase-polymerase chain reaction, chest computed tomography, or rapid antigen test) and the reporting coverage on the period 15th of March - 27th of June was estimated to be rough 70 % of all hospitalized COVID-19 cases [40]. The data gathered during the period March 14th, 2020 until June 12th, 2020 were previously analyzed by Faes et al. [41]. The added value of performing a similar analysis in this study is threefold: 1) To include the patient data gathered in the meantime. 2) To compute the age-stratified mortalities in the cohort and IC hospital wards (*m*_*C*_, *m*_ICU_), as well as the age-stratified recovery time in cohort after a stay in ICU (*d*_ICU,rec_), which were not included by Faes et al. [41]. 3) To obtain age-stratified estimates in nine ten-year age strata as compared to four age strata by Faes et al. [41]. For every patient the following data were provided: 1) age, 2) sex, 3) date of onset of symptoms 4) hospital admission date, 5) hospital discharge date, 6) date of ICU transfer, 7) the number of days spent in ICU, 8) outcome (recovered or deceased). Data from 30 191 patients were excluded from the analysis because one or more of the above entries were missing or because the computed residence times were negative. Patients that came from a nursing home were excluded from the analysis because their inclusion skewed the model predicted number of hospital deaths when the obtained hospitalization parameters were propagated in the model. Thus, in total, data from the remaining 22 136 patients were used (Figure 14). The confidence intervals of the mortalities (*m*_C,ICU_, *m*_C_ and *m*_ICU_) and the distribution between the cohort and IC ward (*c*) were computed using bootstrap resampling. For all hospital residence times, the shape and scale parameters of a Weibull distribution were fitted to the data. To determine if the duration of a cohort or ICU stay differed significantly and to determine if the mortalities in cohort and ICU differed significantly, the non-parametric Mann-Whitney U-test was used. Temporal changes in the estimated hospitalization parameters are not considered in this study. The results of the analysis are presented in Section 3.1.

### 2.3 Social contact model

As previously mentioned, the social behavior of the Belgian population must be translated into a linear combination of the aforementioned pre-pandemic interaction matrices. Mathematically, we must find tangible coefficients so that the linear combination of pre-pandemic interaction matrices, i.e.,

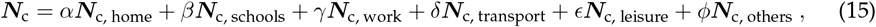

is a good representation of macroscopic social behaviour during the pandemic. Instead of using pre-pandemic contact matrices, modelers would ideally use pandemic contact matrices to build disease models as these are expected to better represent mixing behaviour under lockdown measures. Although these new contact studies under social restrictions will be valuable during future pandemics, such matrices were not available at the start of the pandemic. Hence, our model builds upon pre-pandemic knowledge of social behaviour to make a prediction on pandemic social behavior.

#### Mobility reductions

Google’s Community Mobility Reports (GCMRs) collates data from smartphone users accessing Google applications who allow recording of their *location history* [42]. *The data are categorised into six discrete categories: 1) retail and recreation*, 2) *parks*, 3) *groceries and pharmacies*, 4) *workplaces*, 5) *transport* and 6) *residential* areas. The GCMRs provide the percentage change in activity at each location category compared to that on baseline days before the start of the COVID-19 pandemic (a 5-week period running from 3 January 2020 to 6 February 2020) [15]. The values thus represent the relative change compared to the baseline, and not the absolute number of visitors. The GCMRs are not age-stratified and do not correct for potential underrepresentation of older individuals in the data collection. In our model, the GCMRs for Workplaces, Transit stations, Retail & recreation and Groceries & pharmacy are used as proxies to scale the work (***N***_c, work_), transport (***N***_c, transport_), leisure (***N***_c, leisure_) and other (***N***_c, others_) social contact matrices.

Two surges in COVID-19 cases were observed in Belgium, resulting in two lockdowns (Figure 2). The first lockdown was imposed on March 15th, 2020, and lasted until May 4th, 2020, and involved the closure of schools, bars, clubs, restaurants, all non-essential shops, and a closure of the border to non-essential travel (Table 2). From May 4th, 2020 until July 1st, 2020 the lockdown was gradually lifted. During the first lockdown, schools remained fully closed until May 18th, 2020, and were only re-opened to a very limited extent before the end of the school year on July 1st, 2020. The second lockdown was imposed on October 19th, 2020, and is still ongoing at the time of writing. Schools were closed on November 2nd, 2020, and re-opened on November 16th, 2020. Further, schools were closed during the Christmas holidays from December 18th, 2020 until January 4th, 2021. Universities remained fully closed since October 19th, 2020. Briefly summarized, the first 2020 COVID-19 wave consisted of 1) a rapid surge in cases, 2) a lockdown, and 3) a release of lockdown measures. The second 2020 COVID-19 wave consisted of 1) a rapid surge in cases, 2) a lockdown with schools closed, 3) a lockdown with varying school policies. A more detailed overview of all key events in Belgium during the COVID-19 pandemic is provided in the supplementary materials (Section A.3).

**Figure 2:**
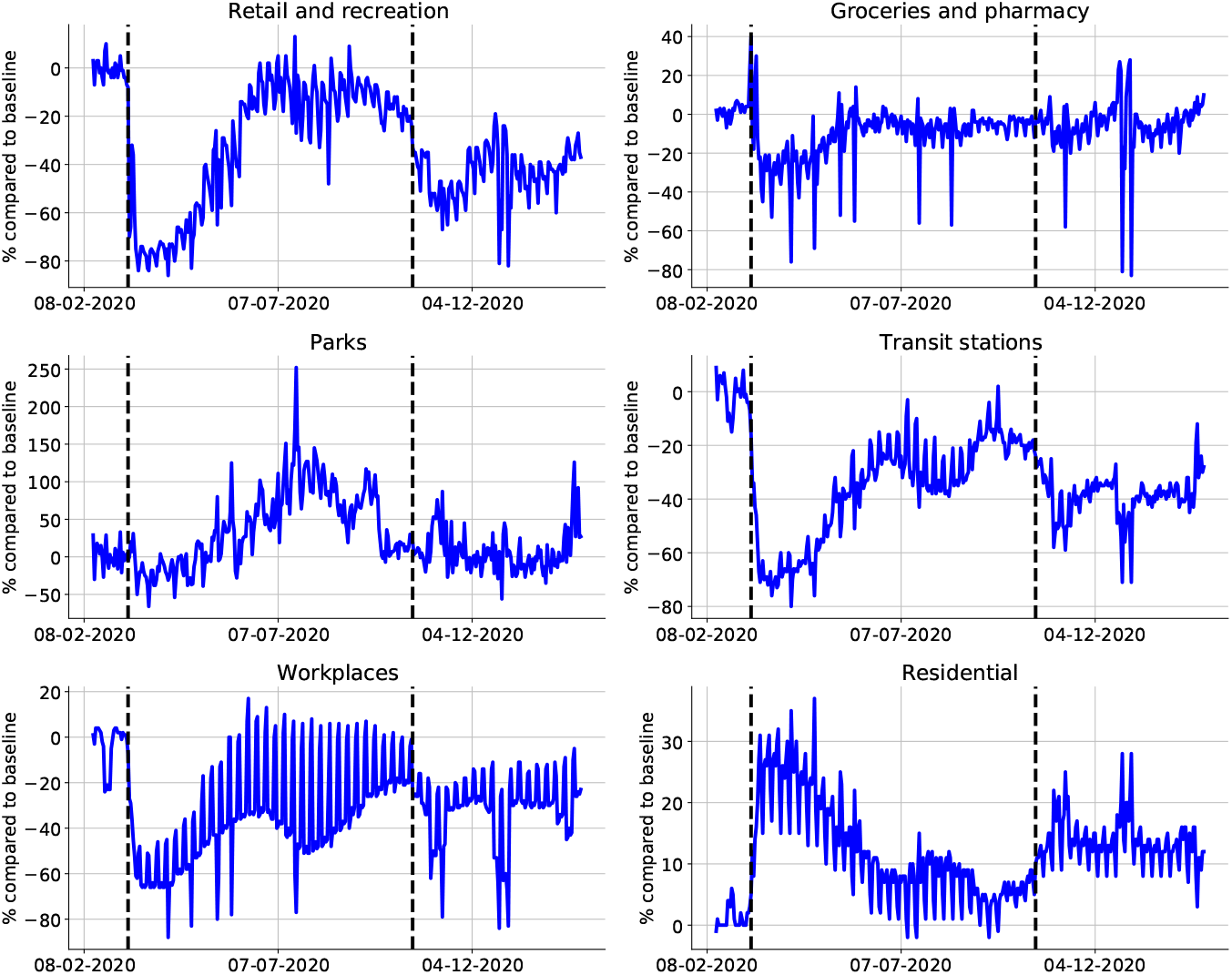
Mobility data extracted from the *Google Community Mobility Reports*. Dashed lines indicate the start of the first lockdown on Friday, March 13th, 2020, and the start of the second lockdown on Monday, October 19th, 2020. Increases in the categories *residential* and *parks* suggest increased activity around the home environment, while increases in the other categories are more indicative of increases in general mobility [43]. The mobility reduction in *workplaces* is used to scale the work interaction matrix, the *retail & recreation* reduction is used to scale the leisure interaction matrix, the *groceries & pharamacy* reduction is used to scale the other interaction matrix, the *transit stations* reduction is used to scale the public transport mobility matrix.

**Table 2:** Dates of key events during the first and second lockdown in Belgium. Google mobility reduction (see Figure 2), computed e as the average reduction between one key event and the next.

| Date | Key event | Details | $G_{\text{work}}$ | $G_{\text{transit}}$ | $G_{\text{r\&r}}$ | $G_{\text{g \& p}}$ | $H_{\text{schools}}$ |
| --- | --- | --- | --- | --- | --- | --- | --- |
| First COVID-19 wave (March - July 2020) |  |  |  |  |  |  |  |
| 15/03/2020 | Lockdown | Closure of schools, bars, clubs and restaurants; Closure of all non-essential shops; Non-essential travel forbidden. [56] | - 56 % | -65 % | -72 % | -26 % | - 100 % |
| 04/05/2020 | Lockdown release phase Ia | Re-opening of industry and B2B services. Re-opening of non-essential retail. Merging of two *social bubbles* allowed [57]. | - 44 % | -54 % | -57 % | -18 % | - 100 % |
| 11/05/2020 | Lockdown release phase Ib | Re-opening of all businesses and shops. Working at home remains the norm where possible. | - 38 % | -45 % | -46 % | -12 % | - 100 % |
| 18/05/2020 | Lockdown release phase IIa | Re-opening of businesses that involve the most human-human contact (f.i. hairdressers). Re-opening of schools for graduating classes in elementary and secondary education [58]. | - 38 % | -39 % | -39 % | -8 % | - 100 % |
| 04/06/2020 | Lockdown release phase III | Re-opening of bars and restaurants. Gatherings up to 10 persons are allowed. | -22 % | -27 % | -15 % | -4 % | - 100 % |
| 01/07/2020 | Lockdown release phase IV | Closure of schools for summer holidays. Gatherings of up to 15 persons are allowed. | -32 % | -27 % | -11 % | -8 % | - 100 % |
| 01/08/2020 | Antwerp Lock-down | The number of infections starts increasing in Antwerp province, where a second lockdown with curfew is imposed [59]. | -28 % | -33 % | -32 % | -6 % | - 100 % |
| Second COVID-19 wave (September 2020 - present) |  |  |  |  |  |  |  |
| 01/09/2020 | End of summer holidays | Opening of elementary and secondary schools. | -18 % | -17 % | -14 % | -5 % | - 0 % |
| 19/10/2020 | Lockdown | Closure of bars and restaurants; Curfew; Strict social restrictions. [60] | -26 % | -31 % | -39 % | -3 % | - 0 % |
| 02/11/2020 | Lockdown | Closure of non-essential stores; Closure of all schools. [61] | -43 % | -48 % | -55 % | -13 % | - 100 % |
| 16/11/2020 | Schools reopen | Elementary and secondary schools reopen. Universities remain closed. | -27 % | -37 % | -44 % | -5 % | - 0 % |
| 12/18/2020 - 04/01/2021 | Christmas holidays | Elementary and secondary schools close. Decrease in work related mobility. | -45 % | -47 % | -42 % | -4 % | - 100 % |
| 04/01/2021 - 15/02/2021 | Period between holidays | Elementary and secondary schools reopen. British variant (501Y.V1) starts spreading [62]. Vaccination campaign in elderly homes starts [63]. | -27 % | -38 % | -43 % | -6 % | - 0 % |

During both lockdowns, mobility increases in the categories *residential* and *parks* were observed (Figure 2). These are indicative of decreased mobility, as these suggest increased activity around the home environment. The other four categories are more indicative of general mobility as they are related to activity around workplaces, retail outlets and use of public transportation [43]. Thus, although the mobility figures indicate people spent more time at home, this does not mean people have more contacts at home (especially under stay-at-home orders). Amplifying the fraction of household contacts under lockdown measures would increase intergenerational mixing of the population under lockdown, which is unrealistic and will lead to overestimations of the hospitalizations. The inability to accurately capture the disease spread in home *bubbles* under lockdown measures is an inherent downside of compartmental epidemiological models. We have thus not scaled the home interaction matrix (***N***_c,home_) with the residential mobility from the GCMRs.

#### Effectivity parameters

During the first lockdown, we estimated that the overall fraction of the social contacts that contributed to SARS-CoV-2 spread, from hereon referred to as the *effectiveness* of the contacts (Ω), was approximately one third of what would be expected based on the GCMRs reductions and the pre-pandemic contacts. Over the course of the first lockdown, work mobility decreased by 56 %, the public transport mobility decreased by 65 %, leisure mobility decreased by 72 % and grocery (others) mobility decreased by 26 % (Table 2). Mathematically,

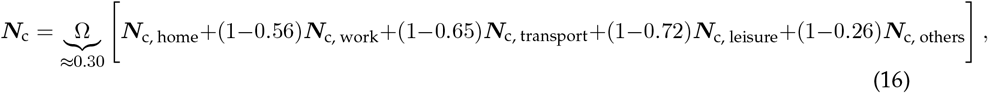

Intuitively, the effectivity of a contacts may not scale linearily with the observed mobility reductions. The net effectivity of the contacts under lockdown measures depends on a combination of the pre-pandemic physical proximity and duration of the contact, the effectivity of preventive measures and on behavioural changes. As an example, the effects of alcohol gel and face masks might be large in the workplace and in grocery stores, but not at home or during leisure activities. To account for different effectivities of contacts in different places, we could introduce one additional parameter per contact matrix, bound between zero and one, and infer its distribution from the available hospitalization data. However, estimating six effectivity parameters was unfeasible because of identifiability issues. We determined that the effectivity parameters of public transport and other places could not be identified. This is most likely because very little contacts are made in those places [44]. Consequently, the effectivity parameters of public tranport, other places and leisure contacts were aggregated to reduce the number of effectivity parameters from six to four. Finally, the linear combination of interaction matrices used to represent social contact under lockdown measures is,

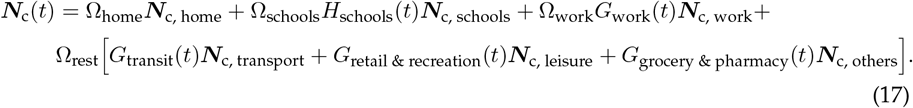

Here, ***N***_c, home_, ***N***_c, schools_, ***N***_c, work_, ***N***_c, transport_, ***N***_c, leisure_ and ***N***_c, others_ denote the pre-pandemic contact matrices at home, in schools, in workplaces, on public transport, during leisure activities and during other activities [14]. *G*_work_, *G*_transit_, *G*_retail & recreation_ and *G*_grocery & pharmacy_ denote the GCMRs mobility reductions in the respective categories and our updated at every timestep in the simulations. *H*_schools_ denotes the fraction of schools opened, as school opening cannot be deduced from the GCMRs. In spite of their limited re-opening on May 18th, 2020, schools are assumed to be closed during the first lockdown. Ω_home_, Ω_schools_, Ω_work_, Ω_rest_ are the effectivity parameters at home, in schools, at work and during leisure, public transport and other activities.

#### Obedience to measures

In reality, compliance to social restrictions is gradual and cannot be modeled using a step-wise change of the social interaction matrix ***N***_***c***_(*t*) (Section 2.1.3). This can be seen upon close inspection of the GCMRs after lockdown measures were taken (Figure 2). Because Google mobility data are updated daily in the model, the effect of gradual mobility changes is inherently included. However, the added value of a social compliance model is to gradually introduce the effects of the effectivity parameters in the model. Further, since the compliance model parameters will be estimated from hospitalization data, the added degrees of freedom aid in obtaining a better model fit to the peak hospitalizations. In our model, we use a delayed ramp to model compliance, i.e.,

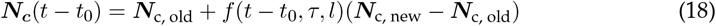

where,

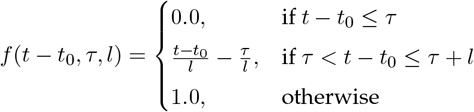

where *τ* is the number of days before measures start having an effect and *l* is the number of additional days after the time delay until full compliance is reached. Both parameters are calibrated to the daily number of hospitalizations in Belgium (Section 2.4). The difference *t* − *t*_0_ denotes the number of days since a change in social policy.

### 2.4 Parameter identification and model predictions

#### Aim of the calibration procedure

To demonstrate the robustness of the social contact model and calibration method, for each of the 2020 COVID-19 waves, we calibrate the model to a minimal dataset and then increase the amount of data used in the calibration procedure to assess if the model can adequately predict future hospitalizations and to assess if the posterior distributions of the effectivity parameters (Ω_*x*_) convergence. For the first COVID-19 epidemic, we calibrate the model using data until April 4th, 2020, and then extend the data range used in the calibration in two-week increments until July 1st, 2020. During the second wave, we calibrate the model until November 7th, 2020, and then extend the calibration to the date of schools re-opening until November 16th, 2020, the date of schools closing for Christmas holidays on December 18th, 2020 and we finally calibrate until February 1st, 2021. By February 1st, 2021, the full impact of school closure and decrease in work mobility during the holiday period is visible in the new hospitalizations. Extending the calibration beyond February 1st, 2021 is out of scope for this study, as the emergence of more contagious strains (B.1.1.7) and the national vaccination campaign need to be included from this point onward (Table 2). As previously mentioned, the effectiveness of contacts in schools cannot be studied during the first COVID-19 wave because schools were only opened to a very limited extent before their final closure on July 1st, 2020.

#### Parameters

The model parameters *R*_0_, *l, τ*, Ω_home_, Ω_schools_, Ω_work_, Ω_rest_ and *ζ* must be calibrated to the available hospitalization data. From Equation 14, the basic reproduction number depends on four model parameters, *β, ω, d*_*a*_ and *a*_*i*_. We calibrate *β* and *d*_*a*_ to hospitalization data. *ω* is ommitted from the calibration because an increase in *ω* can be compensated by a decrease in *d*_*a*_. The subclinical fraction *a*_*i*_ is ommitted because it is an age-stratified parameter consisting of nine values, which renders its calibration computationally unfeasable. The calibration of *β* and *d*_*a*_ should allow sufficient degrees of freedom to obtain a robust estimate of the basic reproduction number without increasing the demand for computational resources too much. In total, nine parameters must be calibrated to hospitalization data.

#### Data

The calibration procedure aims to obtain a parameter set that leads to a good agreement between the model predictions and the observed data. We calibrate all parameters except the seroreversion rate (*ζ*) to the time-series of daily new hospitalizations (*H*_in_), which are available for download at https://epistat.sciensano.be/Data. The seroreversion rate is estimated using five serological measurements from Herzog et al. [39] and eight serological measurements from Sciensano, spanning a period from March 30th, 2020 until July 7th, 2020. For the sake of computational efficacy, the model is first calibrated to the first COVID-19 wave in Belgium, then, the model states on September 1st, 2020 are used as the initial condition to initiate the calibration of the second COVID-19 wave. In this way, the calibration procedure is split between the first COVID-19 wave from March 15th, 2020 until July 1st, 2020, and the second COVID-19 wave from September 1st, 2020 until February 1st, 2021.

#### Statistical model

Rather than using the method of least squares, we assume the data are independent and identically distributed (i.d.d.) sequences of poisson variables. The resulting log-likelihood function is,

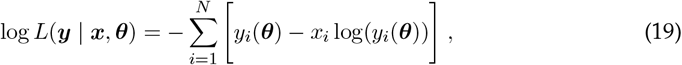

where the vector of parameters, ***θ***, that maximizes the log-likelihood function must be found. In Equation 19, ***y*** denotes the model prediction, ***x*** denotes the timeseries of data and *N* represents the number of datapoints.

#### Calibration procedure

The fitting procedure is performed in two steps. Maximising the result of Equation 19 is computationally demanding and suffers from the presence of local maxima. We thus need an efficient way to scan through the nine-dimensional parameter space ***θ*** = {*β, d*_*a*_, …, Ω_rest_, *ζ*}. A good technique to initially broadly identify the region where the global maximum is situated is *Particle Swarm Optimisation* (PSO) [45]. When a region of interest has been identified, we use the maximum-likelihood estimates as initial values for the ensemble sampler for Markov Chain Monte Carlo (MCMC) proposed by Goodman and Weare [16]. For all parameters, uniform prior distributions were used.

### 2.5 Effects of non-pharamaceutical interventions

To better compare the effects of mobility changes on the daily number of new hospitalizations, we compute the relative share of contacts and the effective reproduction number (*R*_*e*_) at home, in schools, in workplaces and for the combination of leisure, public transport and other contacts. The number of effective contacts in the aforementioned places at time *t* are equal to,

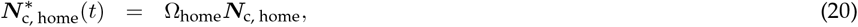

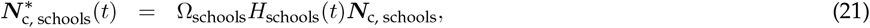

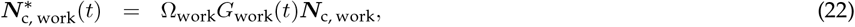

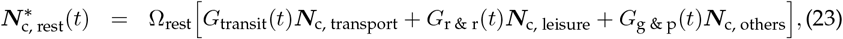

where 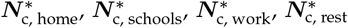 denote the number of effective contacts at home, in schools, at work or for the sum of leisure, public transport and other contacts. The relative share of contacts in location *x* and for age group *i* is computed as,

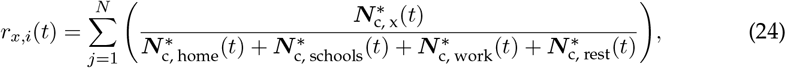

The effective reproduction number for age goup *i*, in place *x* and at time *t* is computed as,

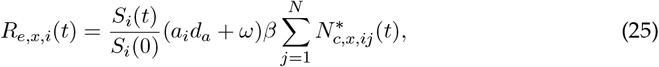

Finally, the population average effective reproduction number in place *x*, and the population average relative share of contacts in location *x*, are computed as the weighted average over all age groups using the demographics listed in Table 1.

## 3 Results

### 3.1 Analysis of hospital surveillance data

The average time from symptom onset to hospitalization is 6.4 days (IQR 2.0 - 8.0 days). Of the 22 136 hospitalized patients, 3 624 patients (16.2 %) required intensive care at some point during their stay and 18 512 (83.8 %) remained in cohort. The overall mortality in the hospital is 21.4 %, the mortality in cohort was significantly lower than the mortality in ICU (16.6 % vs. 46.3 %, *p <* 0.001). One patient under 20 years old has died from COVID-19, mortality is generally low for young patients and increases with older age (Figure 16 and Table 4 of the supplementary materials). The average length of the stay in a cohort ward was 11.0 days (IQR: 4.0 - 13.0 days) and the average length of an ICU stay was 13.6 days (IQR: 4.0 - 19.0 days) (*p <* 0.001). The average cohort stay was 10.8 days (IQR: 4.0 - 12.0 days) if the patient had recovered and 11.8 days (IQR: 4.0 - 14.0 days) if the patient had died (*p <* 0.001). The average ICU stay was 12.0 days (IQR: 3.0 - 15.0 days) if the patient had recovered and 15.2 days (IQR: 5.0 - 21.0 days) if the patient had died (*p <* 0.001). Patients recovering from their ICU stay spend 11.2 additional days (IQR: 4.0 - 13.0 days) in cohort for a recovery and observation stay. Residence times in cohort are shorter than residence times in ICU. In both wards, deceased patients had longer stays than recovered patients (Figure 15 and Table 5 of the supplementary materials). Residence times in cohort and ICU increase with the patient’s age, the same goes for the length of a recovery stay after a stay in ICU. For example, a 20-30 year old patient is expected to spend 6.3 days (IQR: 2.0 - 7.0 days) in cohort while a 70-80 year old patient is expected to spend 12.6 days in cohort (IQR: 5.0 - 14.0 days) (Table 5).

**Table 3:** Results of the time-lagged cross-correlation between the number of cases in the age groups [0 − 20[, [20 − 60[ and [60 − ∞[. Data from November 2nd, 2020 until February 1st 2020 were used in the analysis, which is equal to the daterange range shown in Figure 4.

| Age group (years) | Time-lag (days) | Covariance (-) |
| --- | --- | --- |
| $[0-20[$ vs. $[20-60[$ | -9 | 0.72 |
| $[0-20[$ vs. $[60-\infty[$ | -13 | 0.70 |
| $[20-60[$ vs. $[60-\infty[$ | 0 | 0.98 |

**Table 4:**
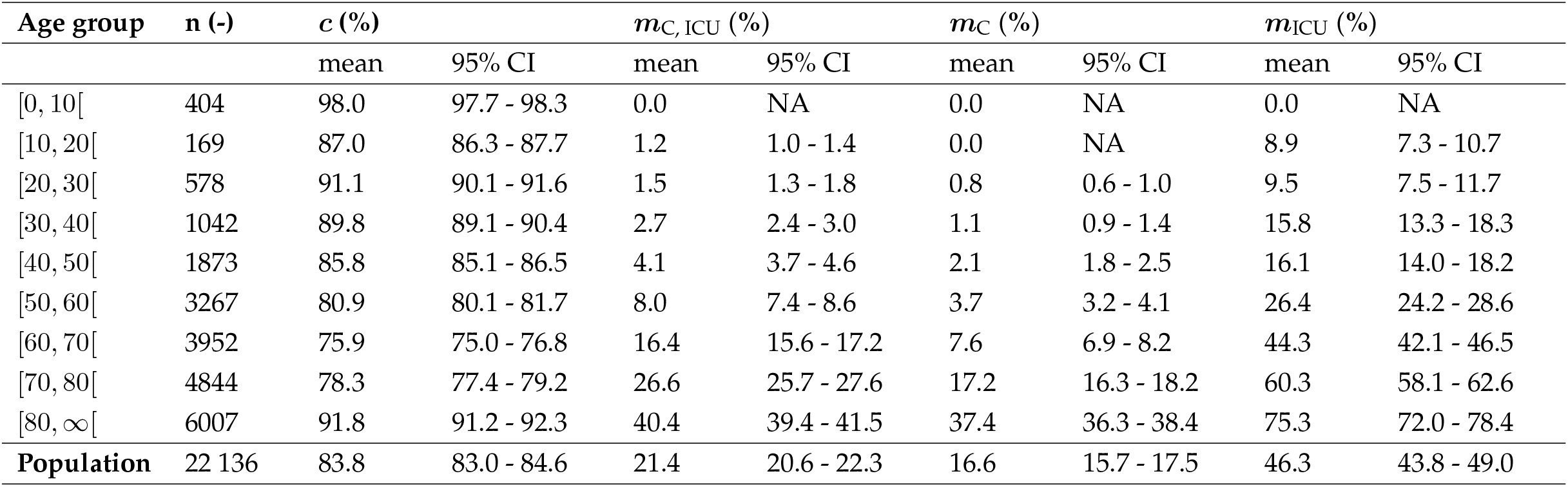
Computed fraction of hospitalized patients remaining in cohort and not transferring to ICU (***c***), pooled mortality in cohort CU (***m***_C, ICU_), mortality in cohort and ICU (***m***_C_) and mortality in ICU (***m***_ICU_) per ten-year age strata. Estimates obtained by bootstrap resampling from the Belgian COVID-19 clincial surveillance on hospitalizations by Van Goethem et al. [40].

| Age group | n (-) | $c$ (%) | | $m_{C,ICU}$ (%) | | $m_C$ (%) | | $m_{ICU}$ (%) | |
| --- | --- | --- | --- | --- | --- | --- | --- | --- | --- |
|  |  | mean | 95% CI | mean | 95% CI | mean | 95% CI | mean | 95% CI |
| [0, 10[ | 404 | 98.0 | 97.7 - 98.3 | 0.0 | NA | 0.0 | NA | 0.0 | NA |
| [10, 20[ | 169 | 87.0 | 86.3 - 87.7 | 1.2 | 1.0 - 1.4 | 0.0 | NA | 8.9 | 7.3 - 10.7 |
| [20, 30[ | 578 | 91.1 | 90.1 - 91.6 | 1.5 | 1.3 - 1.8 | 0.8 | 0.6 - 1.0 | 9.5 | 7.5 - 11.7 |
| [30, 40[ | 1042 | 89.8 | 89.1 - 90.4 | 2.7 | 2.4 - 3.0 | 1.1 | 0.9 - 1.4 | 15.8 | 13.3 - 18.3 |
| [40, 50[ | 1873 | 85.8 | 85.1 - 86.5 | 4.1 | 3.7 - 4.6 | 2.1 | 1.8 - 2.5 | 16.1 | 14.0 - 18.2 |
| [50, 60[ | 3267 | 80.9 | 80.1 - 81.7 | 8.0 | 7.4 - 8.6 | 3.7 | 3.2 - 4.1 | 26.4 | 24.2 - 28.6 |
| [60, 70[ | 3952 | 75.9 | 75.0 - 76.8 | 16.4 | 15.6 - 17.2 | 7.6 | 6.9 - 8.2 | 44.3 | 42.1 - 46.5 |
| [70, 80[ | 4844 | 78.3 | 77.4 - 79.2 | 26.6 | 25.7 - 27.6 | 17.2 | 16.3 - 18.2 | 60.3 | 58.1 - 62.6 |
| [80, ∞[ | 6007 | 91.8 | 91.2 - 92.3 | 40.4 | 39.4 - 41.5 | 37.4 | 36.3 - 38.4 | 75.3 | 72.0 - 78.4 |
| Population | 22 136 | 83.8 | 83.0 - 84.6 | 21.4 | 20.6 - 22.3 | 16.6 | 15.7 - 17.5 | 46.3 | 43.8 - 49.0 |

**Table 5:** Hospital residence time in cohort, irregardless of COVID-19 outcome (***d***_***C***_), residence time in cohort, in case of recovery **(*d***_***C***,***R***_), residence time in cohort, in case of death (***d***_***C***,***D***_). Hospital residence time in IC, irregardless of COVID-19 outcome (***d***_***ICU***_), residence time in IC, in case of recovery (***d***_***ICU***,***R***_), residence time in IC, in case of death (***d***_***ICU***,***D***_) per ten-year age strata. Scale and shape parameters of Weibull distribution fitted to the residence time data. Estimates obtained by analyzing a subset of data from the belgian COVID-19 clincial surveillance on hospitalizations by Van Goethem et al. [40].

| Age group | $d_C$ (days) | | | | $d_{C,R}$ (days) | | | | $d_{C,D}$ (days) | | | |
| --- | --- | --- | --- | --- | --- | --- | --- | --- | --- | --- | --- | --- |
|  | mean | IQR | scale | shape | mean | IQR | scale | shape | mean | IQR | scale | shape |
| [0, 10[ | 3.4 | 2.0 - 4.0 | 3.66 | 1.22 | 3.4 | 2.0 - 4.0 | 3.66 | 1.22 | NA | NA | NA | NA |
| [10, 20[ | 6.3 | 2.0 - 7.0 | 5.64 | 0.85 | 6.3 | 2.0 - 7.0 | 5.64 | 0.85 | NA | NA | NA | NA |
| [20, 30[ | 4.9 | 2.0 - 5.0 | 4.86 | 0.98 | 4.9 | 2.0 - 5.0 | 4.86 | 0.98 | 5.0 | 3.5 - 6.0 | 5.67 | 2.10 |
| [30, 40[ | 5.5 | 3.0 - 6.0 | 5.77 | 1.12 | 5.5 | 3.0 - 6.0 | 5.77 | 1.12 | 6.1 | 2.0 - 11.0 | 6.38 | 1.13 |
| [40, 50[ | 6.3 | 3.0 - 8.0 | 6.81 | 1.22 | 6.3 | 3.0 - 8.0 | 6.81 | 1.22 | 6.8 | 2.3 - 8.8 | 6.94 | 1.03 |
| [50, 60[ | 7.6 | 4.0 - 9.0 | 8.12 | 1.16 | 7.6 | 4.0 - 9.0 | 8.07 | 1.17 | 9.1 | 3.0 - 10.0 | 9.16 | 1.01 |
| [60, 70[ | 10.0 | 4.0 - 11.0 | 10.32 | 1.08 | 9.9 | 4.0 - 11.0 | 10.31 | 1.10 | 11.2 | 3.0 - 14.0 | 10.32 | 0.86 |
| [70, 80[ | 12.6 | 5.0 - 14.0 | 13.11 | 1.10 | 12.6 | 5.0 - 14.0 | 13.24 | 1.13 | 12.6 | 4.0 - 13.0 | 12.42 | 0.97 |
| [80, ∞[ | 15.6 | 6.0 - 19.0 | 16.37 | 1.13 | 17.8 | 8.0 - 22.0 | 19.1 | 1.21 | 11.9 | 4.0 - 15.0 | 12.21 | 1.06 |
| **Population** | 11.0 | 4.0 - 13.0 | 9.09 | 1.21 | 10.8 | 4.0 - 12.0 | 8.72 | 1.24 | 11.8 | 4.0 - 14.0 | 10.97 | 1.08 |
| Age group | $d_{ICU}$ (days) | | | | $d_{ICU,R}$ (days) | | | | $d_{ICU,D}$ (days) | | | |
|  | mean | IQR | scale | shape | mean | IQR | scale | shape | mean | IQR | scale | shape |
| [0, 10[ | 6.0 | 2.0 - 8.3 | 6.40 | 1.19 | 6.7 | 2.0 - 8.5 | 7.37 | 1.37 | NA | NA | NA | NA |
| [10, 20[ | 4.9 | 2.0 - 5.0 | 5.26 | 1.25 | 4.0 | 2.0 - 5.0 | 4.44 | 1.43 | 16.0 | NA | NA | NA |
| [20, 30[ | 9.6 | 2.0 - 10.0 | 8.86 | 0.87 | 8.9 | 2.0 - 10.0 | 8.34 | 0.89 | 18.0 | 4.5 - 25.5 | 16.97 | 0.89 |
| [30, 40[ | 10.1 | 2.0 - 13.3 | 11.08 | 1.00 | 9.4 | 2.0 - 11.0 | 8.72 | 0.87 | 14.0 | 5.0 - 20.0 | 14.86 | 1.20 |
| [40, 50[ | 11.3 | 3.0 - 14.0 | 12.75 | 1.00 | 10.6 | 3.0 - 12.0 | 10.34 | 0.95 | 15.1 | 4.5 - 21.0 | 16.38 | 1.30 |
| [50, 60[ | 14.1 | 5.0 - 19.0 | 1.05 | 1.00 | 11.7 | 4.0 - 15.8 | 12.02 | 1.08 | 19.7 | 8.5 - 27.0 | 20.60 | 1.16 |
| [60, 70[ | 14.7 | 5.0 - 21.0 | 1.05 | 1.00 | 13.2 | 4.0 - 17.0 | 13.00 | 0.97 | 16.5 | 6.0 - 23.0 | 17.52 | 1.21 |
| [70, 80[ | 15.0 | 5.0 - 21.0 | 1.05 | 1.00 | 14.6 | 4.0 - 21.0 | 14.47 | 0.98 | 15.2 | 6.0 - 21.0 | 15.83 | 1.12 |
| [80, ∞[ | 10.8 | 3.0 - 14.0 | 12.58 | 1.00 | 7.9 | 2.0 - 9.0 | 7.54 | 0.92 | 11.7 | 3.0 - 15.0 | 11.25 | 0.92 |
| **Population** | 13.6 | 4.0 - 19.0 | 11.41 | 1.21 | 12.0 | 3.0 - 15.0 | 12.32 | 0.98 | 15.2 | 5.0 - 21.0 | 13.77 | 1.10 |

**Table 6:** Hospital residence time for a recovery stay in cohort, after a stay in ICU (***d***_ICU,rec_), time from symptom onset to hospitalization (***d***_hospital_) per ten-year age strata. Scale and shape parameters of Weibull distribution fitted to the residence time data. Estimates obtained by analyzing a subset of data from the Belgian COVID-19 clincial surveillance on hospitalizations by Van Goethem et al. [40].

| Age group | $d_{\text{ICU,rec}}$ (days) | | | | $d_{\text{hosp}}$ (days) | | | |
| --- | --- | --- | --- | --- | --- | --- | --- | --- |
|  | mean | IQR | scale | shape | mean | IQR | scale | shape |
| [0, 10[ | 9.9 | 0.5 - 3.0 | 3.18 | 0.40 | 2.2 | 0.0 - 2.0 | 0.86 | 0.43 |
| [10, 20[ | 3.4 | 3.0 - 4.0 | 2.99 | 0.70 | 5.6 | 2.0 - 6.0 | 4.69 | 0.73 |
| [20, 30[ | 8.4 | 3.0 - 10.8 | 8.18 | 0.94 | 6.0 | 2.0 - 7.0 | 5.33 | 0.75 |
| [30, 40[ | 6.6 | 2.0 - 7.0 | 5.88 | 0.80 | 6.7 | 3.0 - 9.0 | 6.78 | 1.02 |
| [40, 50[ | 8.2 | 3.0 - 8.0 | 7.97 | 0.94 | 7.4 | 4.0 - 9.0 | 7.69 | 1.14 |
| [50, 60[ | 10.1 | 4.0 - 11.0 | 10.02 | 0.99 | 7.5 | 4.0 - 10.0 | 7.73 | 1.08 |
| [60, 70[ | 11.5 | 4.0 - 14.0 | 11.58 | 1.01 | 6.9 | 3.0 - 9.0 | 6.87 | 0.97 |
| [70, 80[ | 15.2 | 6.0 - 20.0 | 15.29 | 1.02 | 6.6 | 2.0 - 8.0 | 5.72 | 0.75 |
| [80, ∞[ | 13.3 | 6.0 - 16.0 | 14.11 | 1.23 | 5.0 | 1.0 - 7.0 | 3.62 | 0.59 |
| **Population** | 11.2 | 4.0 - 13.0 | 8.39 | 1.40 | 6.4 | 2.0 - 8.0 | 10.11 | 0.63 |

### 3.2 Model calibration

The population average basic reproduction number was computed as *R*_0_ = 4.16 (IQR: 3.90 - 4.39) for the first 2020 COVID-19 wave and as *R*_0_ = 3.69 (IQR: 3.64 - 3.75) for the second 2020 COVID-19 wave. Large differences in the basic reproduction number exist between the different age groups (Figure 3). It is clear that the youths and working-aged population drive the SARS-CoV-2 pandemic while people of ages 70 or above can hardly sustain a SARS-CoV-2 pandemic amongst themselves, this is mainly because elderly individuals have limited social interactions (Figure 3). Still, these individuals make up roughly 35 % of all hospitalizations. The biggest risk group are the individuals aged 50 to 70, which make up roughly 50 % of the expected hospitalizations. The high expected fraction of hospitalizations in this age group is due to a trade-off between social contact and hospitalization risk. These individuals have plenty of social contact and at the same time, have a high propensity to hospitalization.

**Figure 3:**
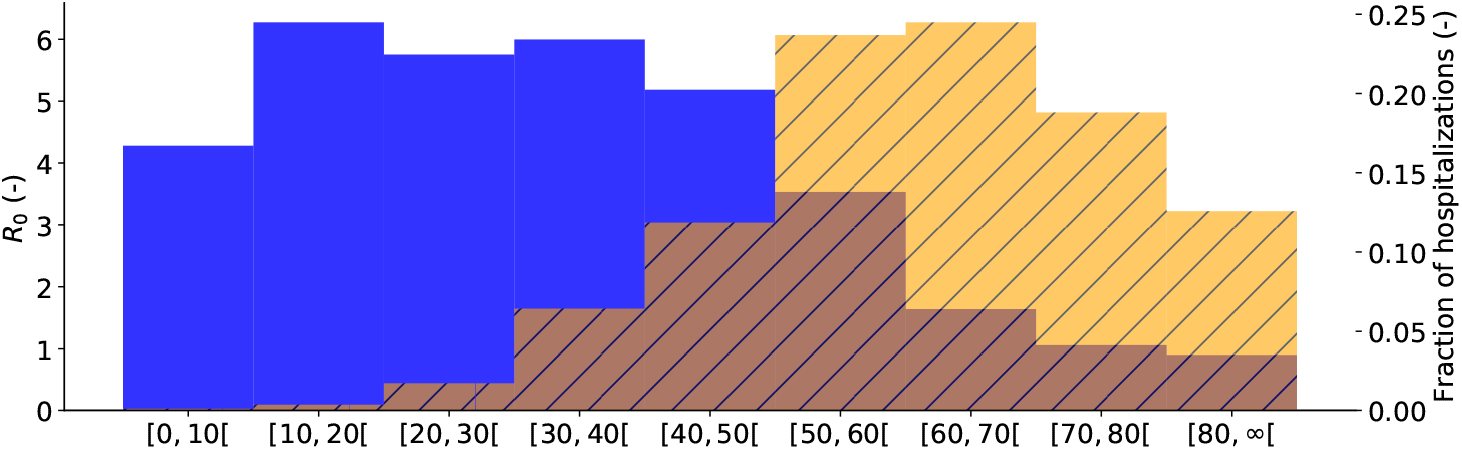
Basic reproduction number per age group (*R*_0,*i*_), for Belgium (blue). Expected fraction of the total Belgian hospitalizations during the first COVID-19 wave, as predicted by the model, from March 15th, 2020 until July 1st, 2020 in age group *i* (orange, striped). Youths and working-aged population drive the pandemic, while the senior population is mostly in need of hospital care.

**Figure 4:**
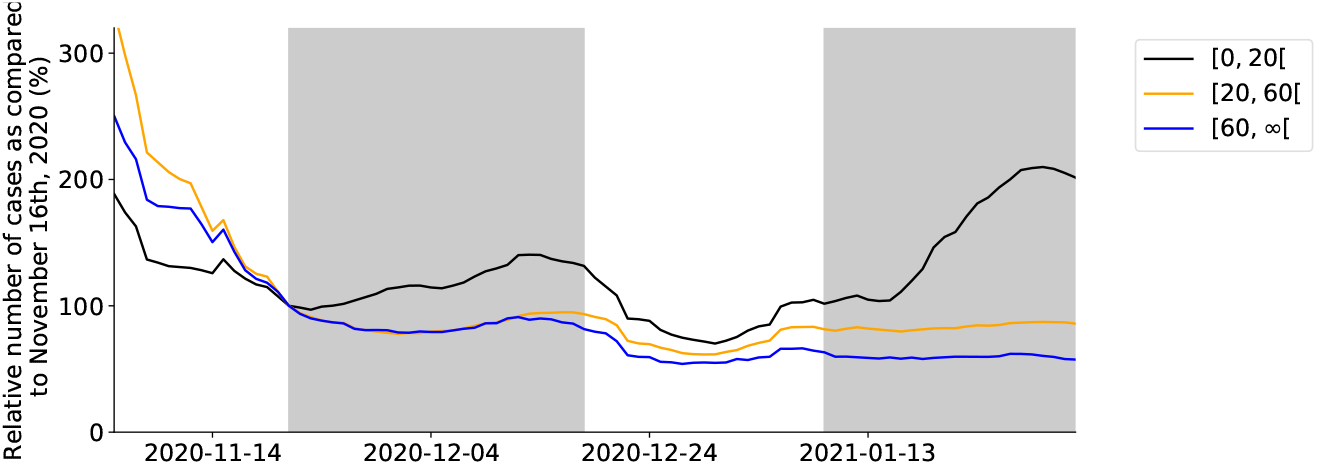
Relative number of confirmed cases in youths, the working population and the senior population during the period November 2nd, 2020 until February 1st, 2021, as compared to the number of confirmed cases in each group on November 16th, 2020. The grey shade is used to indicate schools were open.

Compliance to social measures was similar for both 2020 COVID-19 waves, with an average delay of 0.22 (IQR: 0.07-0.31) and 0.39 (IQR: 0.20 - 0.52) days, and a time to reach full compliance to measures of 9.17 (IQR: 8.89 - 9.50) and 6.94 (IQR: 6.71 - 7.18) days respectively. Using the serological datasets by Herzog et al. [39] and Sciensano, the average time to seroreversion (1*/ζ*) was estimated as 9.2 months (IQR: 7.2 - 12.1 months) (Figure 13). The model was calibrated to the new hospitalizations and serological data, however, to obtain estimates for the total number of patients in Belgian hospitals and the number of deceased patients in Belgian hospitals, the hospitalization parameters computed using the clinical surveillance dataset are propagated in the model using bootstrap sampling. In supplementary figures 5 and 6, the ability of the calibrated model to predict the number of daily hospitalizations, the total number of patients in Belgian hospitals, the total number of deaths in Belgian hospitals, and the seroprevalence in the Belgian population during both 2020 COVID-19 waves are demonstrated. The model’s ability to predict the number of hospital deaths in every age strata is demonstrated in Figure **??**.

Figure 5 summarizes the results of six model calibrations using hospitalization datasets starting on March 15th, 2020 until April 4th, 2020, and subsequently increased in two-week increments. Here, Figure 5 (a) represents the minimal dataset, where the data range used for the calibration was equal to March 15th, 2020 until April 4th, 2020. Opposed is Figure 5f, which uses the maximal dataset, using hospitalization data from March 15th, 2020 until July 1st, 2020. Using the minimal dataset (Figure 5a), the posterior distributions are uninformative and model prediction uncertainty is large. Using additional data from April 15th, 2020 (Figure 5b) onwards, the model captures the observed downward trend in the hospitalization data. Before the release of social restrictions on May 4th, 2020 (Figure 5a-5c), the posterior distributions seem to converge to distributions different from the ones found using the maximal dataset (Figure 5f). However, during the gradual lifting of lockdown restrictions (Figure 5d-5f), the posterior distributions monotonically converge to their final distributions.

**Figure 5:**
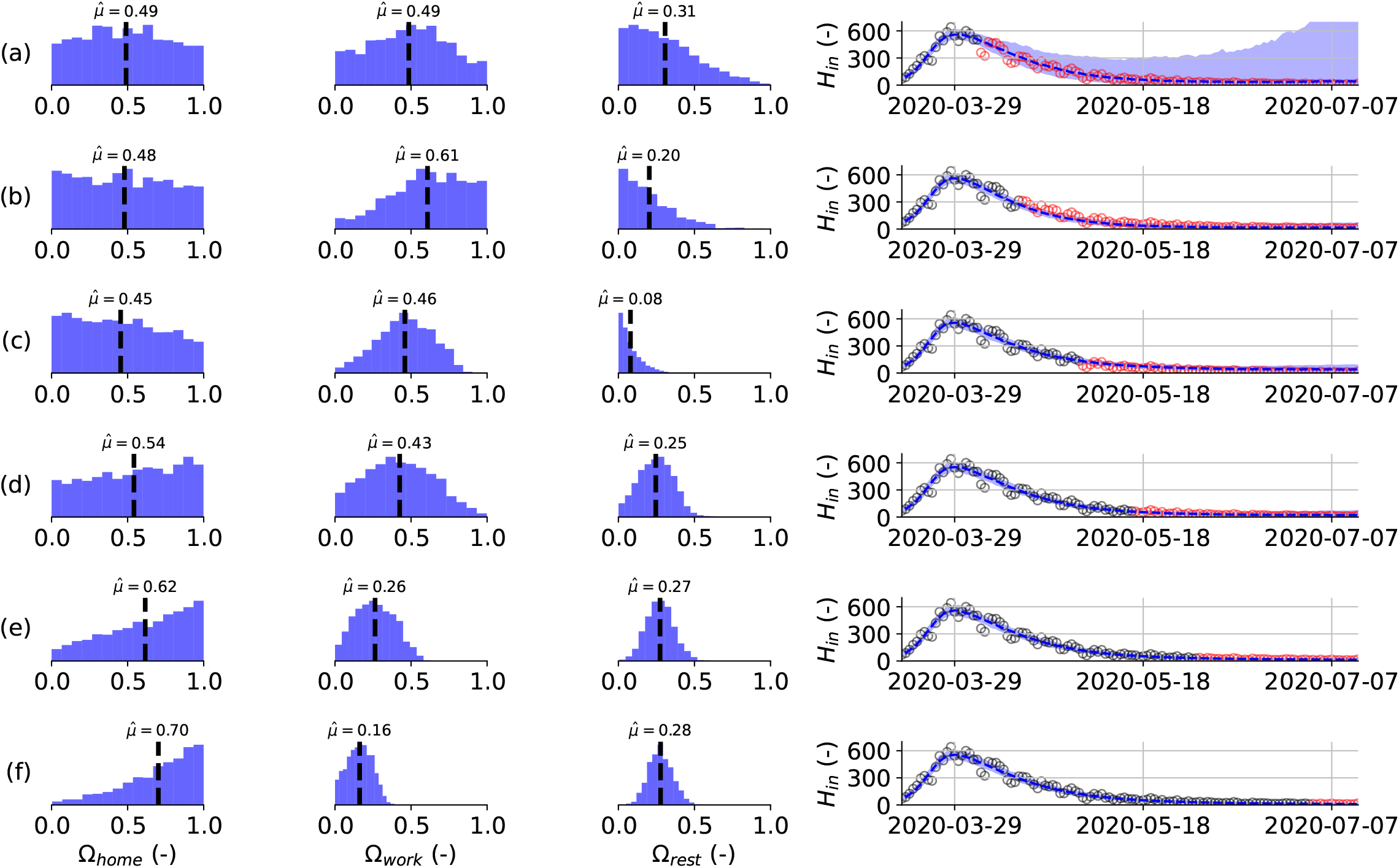
(left) Estimated posterior distributions for the effectivity of a contact at home (Ω_home_), in the workplace (Ω_work_) and for the sum of leisure activities, other activities and public transport (Ω_rest_), (right) together with the resulting model prediction for the daily hospitalizations from March 15th, 2020 until July 14^th^, 2020 (right). The effectivity of school contacts could not be deduced during the first 2020 COVID-19 wave because schools were only re-opened very limited before their final closure on July 1st, 2020. Calibration performed using the daily hospitalizations in Belgium until: (a) 2020-04-04, (b) 2020-04-15, (c) 2020-05-01, (d) 2020-05-01, (e) 2020-06-01 and (f) 2020-07-01. Calibration data in black, validation data in red. Model predictions are accurate in all but the minimal calibration dataset (a). Monotonic convergence of the effectivity parameter posteriors is reached quickly after lockdown release on May 4th, 2020 (d-f).

Similarly, four calibrations on hospitalization datasets of increasing length during the second COVID-19 wave were performed and the results are summarized in Figure 6. Once more, the minimal dataset (Figure 6a), which uses data from September 1st, 2020 until November 7th, 2020 does not result in informative posterior distributions of the effectivity parameters. Uncertainty on the model prediction is large, but the mean model prediction is fairly accurate. As soon as schools are opened on November 16th, 2020, the daily hospitalizations evolve to a plateau. Despite large uncertainty on the model prediction, the emergence of the hospitalization plateau is captured in the uncertainty band, and the model thus provides a starting estimate using the minimal dataset. Although model accuracy has risen, a similar conclusion can be drawn for the calibration using data until schools re-opening on November 16th, 2020. When including data in the hospitalization dataset until schools closure for the Christmas holidays on December 18th, 2020 (Figure 6c), the model correctly attributes the increased transmission to the opening of schools. In Figure 6c, it can be seen that the effectivity parameter for schools is almost equal to the maximum value of one. Although the posteriors of the effectivity parameters still differ significantly from their final distributions, the model provides an accurate prediction for the future evolution of the new hospitalizations during the Christmas holidays and until schools re-opening on January 4th, 2021. From the inference using the maximal dataset (Figure 6d), it is clear that the model attributes high effectivities for contacts at home and in schools.

**Figure 6:**
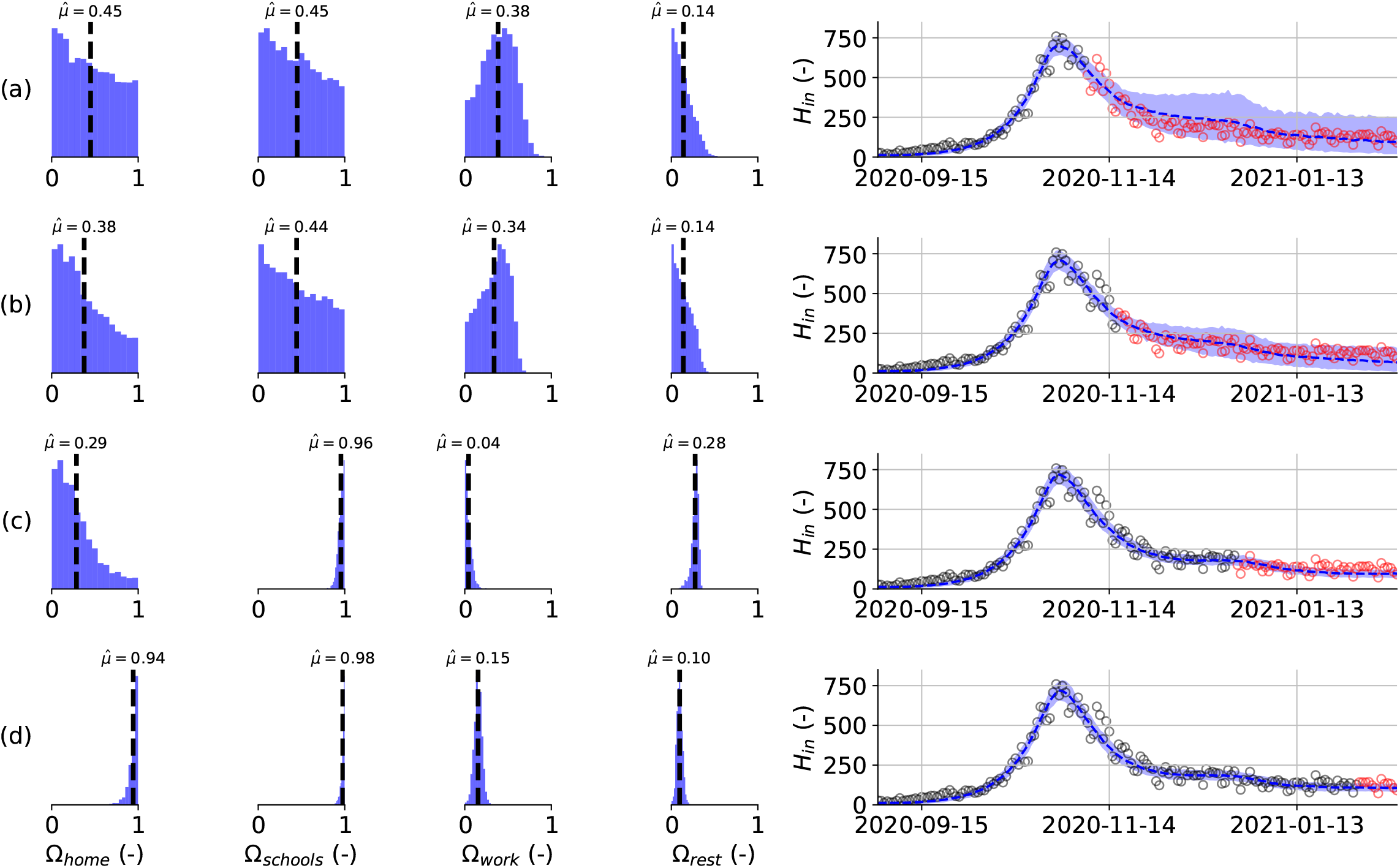
Estimated posterior distributions for the effectivity of a contact at home (Ω_home_), at school (Ω_schools_), in the workplace (Ω_work_) and for the sum of leisure activities, other activities and public transport (left), together with the resulting model prediction for the daily hospitalizations from September 1st, 2020 until February 14th, 2021 (right). Calibration performed using the daily hospitalizations in Belgium until: (a) 2020-11-07, (b) 2020-11-16, (c) 2020-12-18, (d) 2021-02-01. Calibration data in black, validation data in red. Model predictions are accurate for all calibration datasets. Monotonic convergence of the schools effectivity parameter is reached a-posteriori schools re-opening (c).

### 3.3 Effects of non-pharamaceutical interventions

To better compare the effects of non-pharmaceutical interventions between both 2020 COVID-19 waves, we computed the relative share of contacts and the effective reproduction number at home, in schools, in workplaces, and for the sum of leisure, public transport, and other contacts (Figure 7). In this way, we can dissect the force of infection in our model, allowing us to assess the relative impact of contacts made at different locations on SARS-CoV-2 transmission. In pre-pandemic times, leisure and work contacts account for the bulk of total contacts, while under strict lockdown measures (March 15th, 2020 - May 4th, 2020 and October 19th, 2020 - November 16th, 2020), the contacts at home are the main driver of SARS-CoV-2 spread. The effective reproduction number under strict lockdown measures was equal to *R*_*e*_ = 0.67 (IQR: 0.48 - 0.76) for the first COVID-19 epidemic and was equal to *Re* = 0.66 (IQR: 0.61 - 0.69) for the second COVID-19 epidemic. Aside from the interactions at home, leisure contacts had the second most impact during the first COVID-19 wave, with roughly twice the impact of work contacts. When lifting social restrictions from May 4th, 2020 onwards, the relative contribution of home contacts gradually declines, while the contributions of work and leisure become more important. The effective reproduction number gradually increases and approaches the critical value of *R*_*e*_ = 1 by the beginning of summer (average of June, 2020 *R*_*e*_ = 0.91, IQR: 0.77 - 1.00).

**Figure 7:**
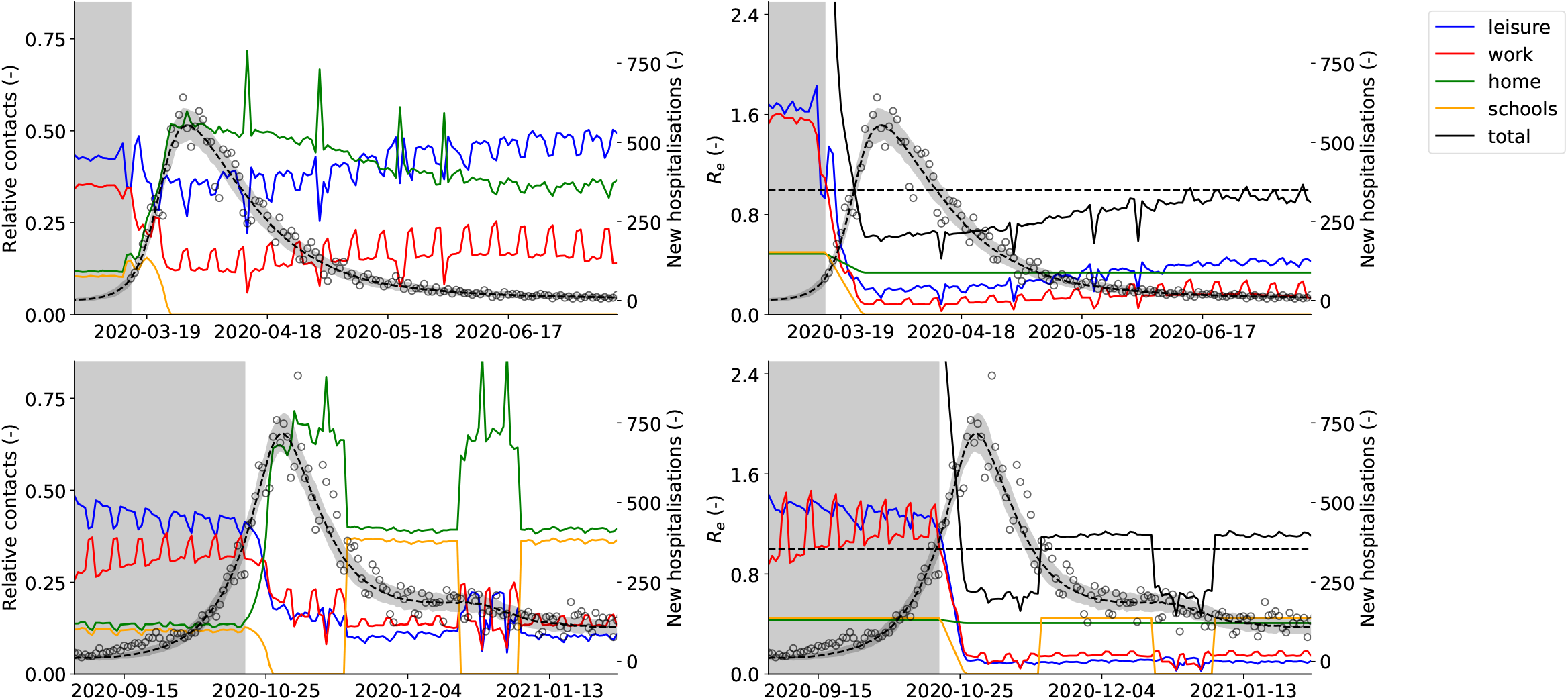
(First column) Relative share of contacts at home, in the workplace, in schools and for the sum of leisure activities, (Second column) effective reproduction number (*R*_*e*_) at home, in the workplace, in schools and for the sum of leisure activities, other activities and public transport. The right axis denotes the predicted number of daily Belgian hospitalizations. The first row depicts the first COVID-19 wave in Belgium, from March 15th, 2020 until July 14th, 2020, while the second row depicts the second COVID-19 wave in Belgium, from September 1st, 2020 until February 1st, 2020. Mean and 95 % confidence interval of 1000 model realisations. The background is shaded grey before lockdown measures were taken. During both lockdowns, home interactions have the largest share of effective contacts. During lockdown release, the relative importance of work and leisure contacts start increasing. Schools opening and closing has a large impact on the effective reproduction number, and can end a decreasing trend in hospitalizations.

As soon as schools are re-opened on November 16th, 2020, a plateau in the daily number of hospitalizations emerges (Figure 7). There were no other major policy changes around this time, except schools re-opening. Our model deduces this correlation by inferring posterior values of the effectivity of contacts in schools close to one, meaning school contacts were highly effective for SARS-CoV-2 transmission. Schools have an impact similar to the home interactions, with both contributing roughly 40 % to the total number of effective contacts during the second COVID-19 wave. The opening of schools under lockdown can tip the scale, and push the effective reproduction number just above the critical value of *R*_*e*_ = 1. When schools are opened, the effective reproduction number increases from *R*_*e*_ = 0.66±0.04 to *R*_*e*_ = 1.09±0.05, causing a stagnation of the daily hospitalizations. To further validate this result, we extracted the number of laboratory-confirmed cases in youths [0, 20[, the working population [20, 60[ and the senior population [60, ∞[ from the *Belgian Scientific Institute of Public Health* (Sciensano). The time-series were normalized with the number of cases on November 21st, 2020^1^ to allow a better comparison. The number of laboratory-confirmed cases amongst youths starts increasing as soon as schools are opened on November 16th, 2020 (Figure 4). A similar pattern is observed during school closure and re-opening for the Christmas holidays, although it should be noted the relationship is less clear. This is most likely the effect of Christmas and New Year celebrations and returning travelers. The use of a time-lagged cross-correlation revealed a significant lead-relationship between the number of cases in youths and the working population by 9 days, and a leading relationship between the number of cases amongst youths and the senior population by 13 days (Section A.5). This indicates that as schools are reopened, SARS-CoV-2 finds its way through social networks from younger to older individuals, eventually pushing the effective reproduction number above one.

## 4 Discussion

### 4.1 Analysis of hospital surveillance data

We computed hospitalization parameters using data from 22 136 patients in Belgian hospitals. The average time from symptom onset to hospitalization was estimated as 6.4 days. This estimate is in line with the previous estimate for Belgium of 5.7 days by Faes et al. [41], and is in line with estimates for other regions such as 5-9 days for China [38], 4.4 days for Hong Kong, and 5.1 days for the UK [46], especially when the interquartile range of 2.0 - 8.0 days is taken into account. Of the 22 136 hospitalized patients, 3 624 patients (16.2 %) required intensive care at some point during their stay and 18 512 (83.8 %) remained in cohort. The result is slightly lower than the estimate of Wu and McGoogan [47] for China, who estimated that one-quarter of all hospitalized patients require intensive care. It should however be noted that the criteria for ICU admission and release might differ between countries. The ICU admission probabilities and mortalities in cohort and ICU indicate that COVID-19 has a much higher severity in older individuals, which is in line with estimates from other studies [4, 48]. In terms of hospital residence times, our estimates agree well with those made by Faes and colleagues [41]. The average time spent in cohort was estimated as 11.0 days (3.4 -15.6 days for the youngest versus oldest age groups), while the average time spent in ICU was estimated as 13.6 days (6.0 - 10.8 days for the youngest versus oldest age groups). The average time spent in ICU was lower in the 80+ age group (10.8 days) than in the 70-80-year-olds (15.0 days). The residence time estimates are in line with Vekaria et al. [49] who estimated a length of stay in England for COVID-19 patients not admitted to ICU of 8.4 days and for ICU length of stay of 12.4 days. It was previously reported by Faes et al. [41] that the median residence time decreased after the first 2020 COVID-19 wave, however, we chose not to account for temporal changes in the hospital residence times and mortalities. The model predicted total number of patients and number of deaths in Belgian hospitals (Figures 9 and 10) would likely benefit from propagating time-dependent hospitalization parameters in the model. Vandromme et al. [50] previously found that the average hospital residence times in Belgium have decreased between the first and second 2020 COVID-19 waves, which is mainly due to standardization of COVID-19 hospital treatment. In spite, the model predictions are sufficiently accurate to aid policymakers in the decision-making process.

**Figure 8:**
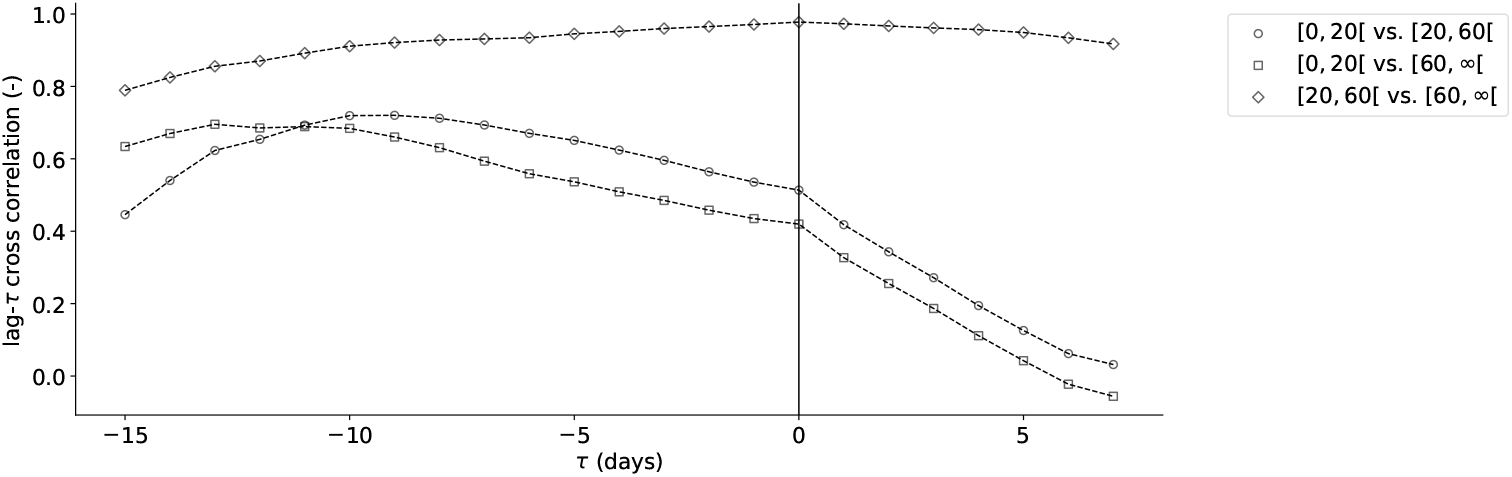
Cross correlation between the number of cases in Belgium in the age groups [0−20[, [20−60[ and [60−∞[, from November 2nd, 2020 until February 1st 2020 in function of the number of days the timeseries are shifted relative to each other (*τ*). The maximum cross correlation is obtained when the series [0 −20[ and [20 − 60[ are shifted -9 days, the maximum cross correlation is obtained when the series [0−20[ and [60−∞[ are shifted -13 days, and the maximum cross correlation is obtained when the series [20 − 60[ and [60 − ∞[ are not shifted.

**Figure 9:**
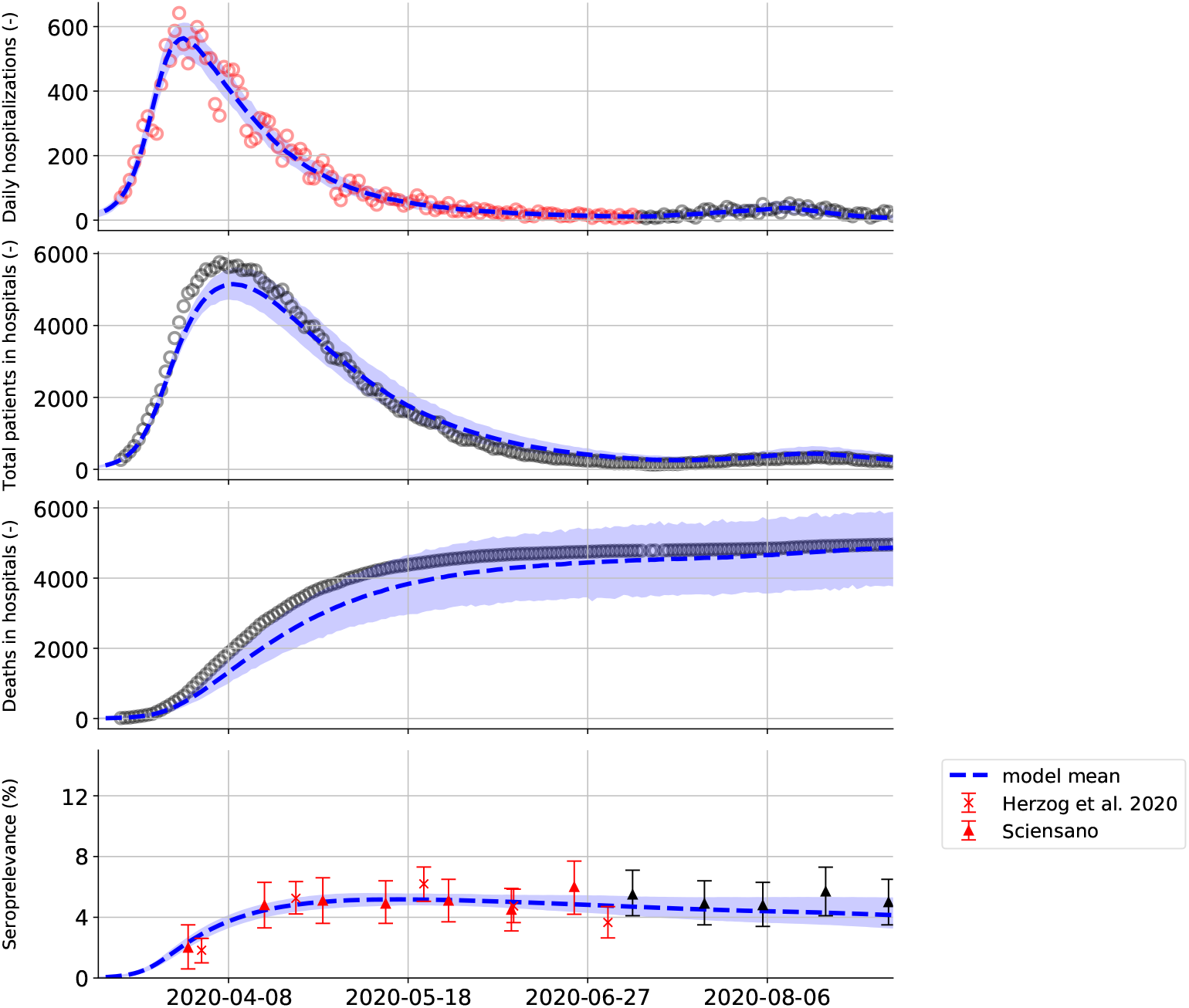
(top to bottom) Model predictions and data during the first COVID-19 wave in Belgium, from March 15th, 2020 until September 1st, 2020: 1) The daily Belgian hospitalizations, 2) the total number of patients in Belgium hospitals, 3) the total number of deceased patients in Belgian hospitals, 4) the seroprelevance in the Belgian population. Mean and 95 % confidence interval of 1000 model realisations. Red datapoints indicate the data was used in the model calibration, black datapoints indicate data was not used in the model calibration. The model is calibrated to the daily Belgian hospitalizations (top), the prediction for the total number of patients in Belgian hospitals and total number of deceased patients in Belgian hospitals are obtained by propagating the age-stratified mortalities (***m***_C_ and ***m***_ICU_), age-stratified distributions between cohort and ICU (***c***) and the residence time distributions derived from the hospital dataset in the model (***d***_C,R_, ***d***_C,ICU_, ***d***_ICU,R_, ***d***_ICU,D_) (see Table 4 and 5).

**Figure 10:**
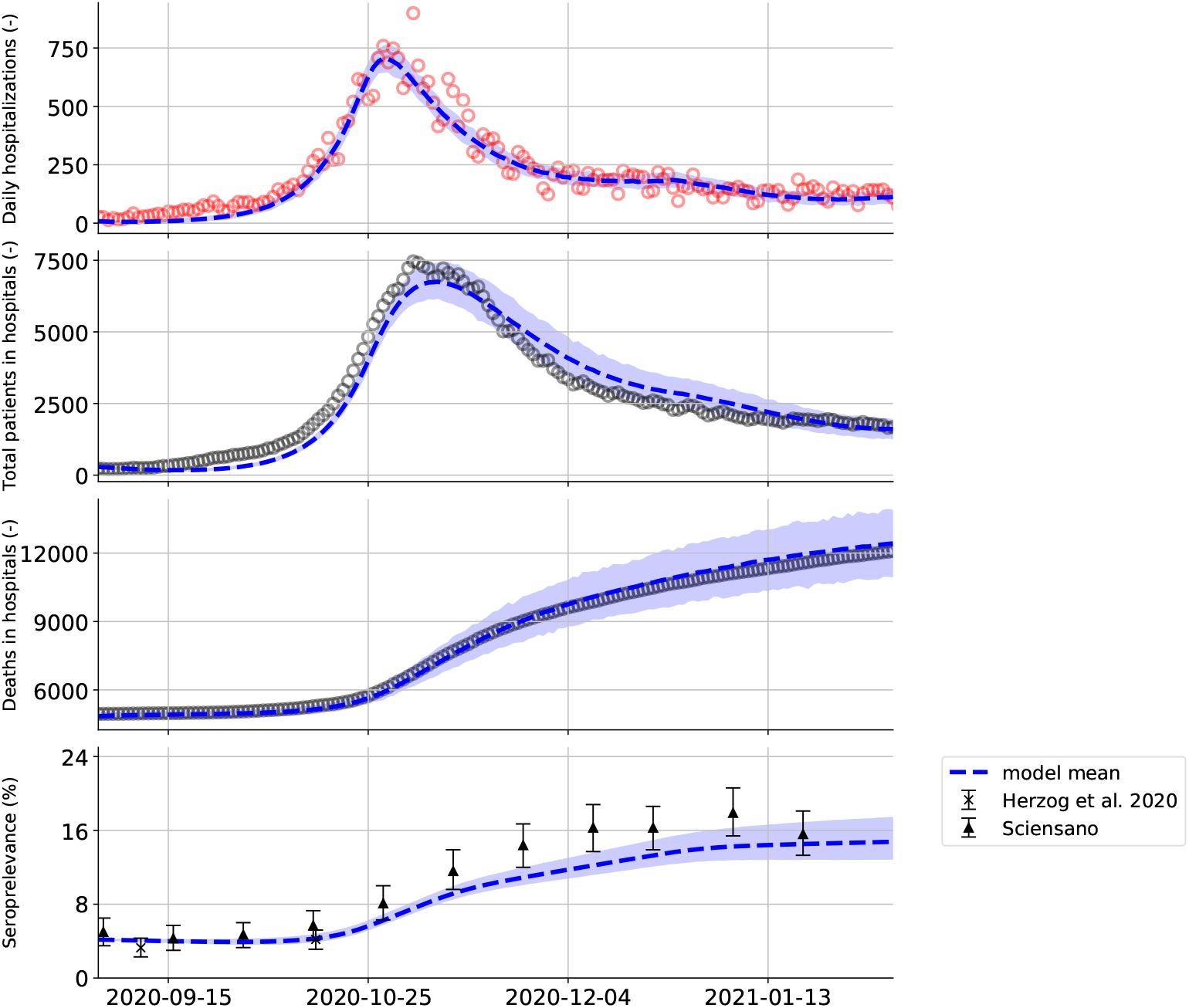
(top to bottom) Model predictions and data during the second COVID-19 wave in Belgium, from September 1st, 2020 until February 1st, 2021: The daily Belgian hospitalizations, 2) the total number of patients in Belgium hospitals, 3) the total number of deceased patients in Belgian hospitals, 4) the seroprelevance in the Belgian population. Mean and 95 % confidence interval of 1000 model realisations. Red datapoints indicate the data was used in the model calibration, black datapoints indicate data was not used in the model calibration. The model is calibrated to the daily Belgian hospitalizations (top), the prediction for the total number of patients in Belgian hospitals and total number of deceased patients in Belgian hospitals are obtained by propagating the age-stratified mortalities (***m***_C_ and ***m***_ICU_), age-stratified distributions between cohort and ICU (***c***) and the residence time distributions derived from the hospital dataset in the model (***d***_C,R_, ***d***_C,ICU_, ***d***_ICU,R_, ***d***_ICU,D_) (see Table 4 and 5).

**Figure 11:**
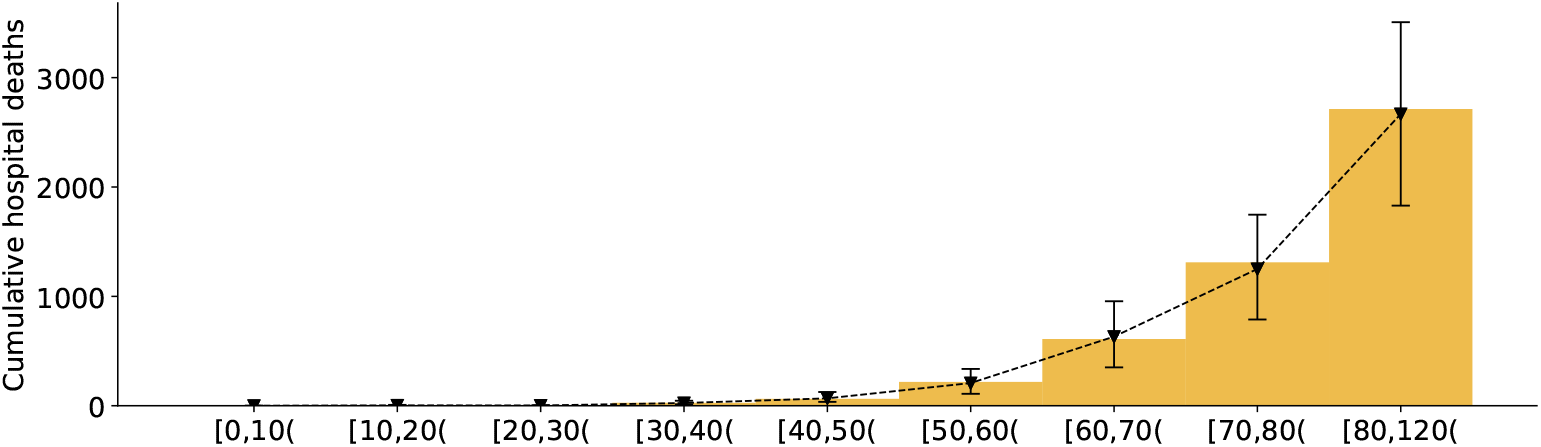
Cumulative deaths in Belgian hospitals per ten-year age strata. For the first Belgian 2020 COVID-19 wave, from March 1st, 2020 until September 1st, 2020. Yellow bars represent the data collected by Sciensano, inverted triangles represent the model prediction mean with 95 % confidence interval.

**Figure 12:**
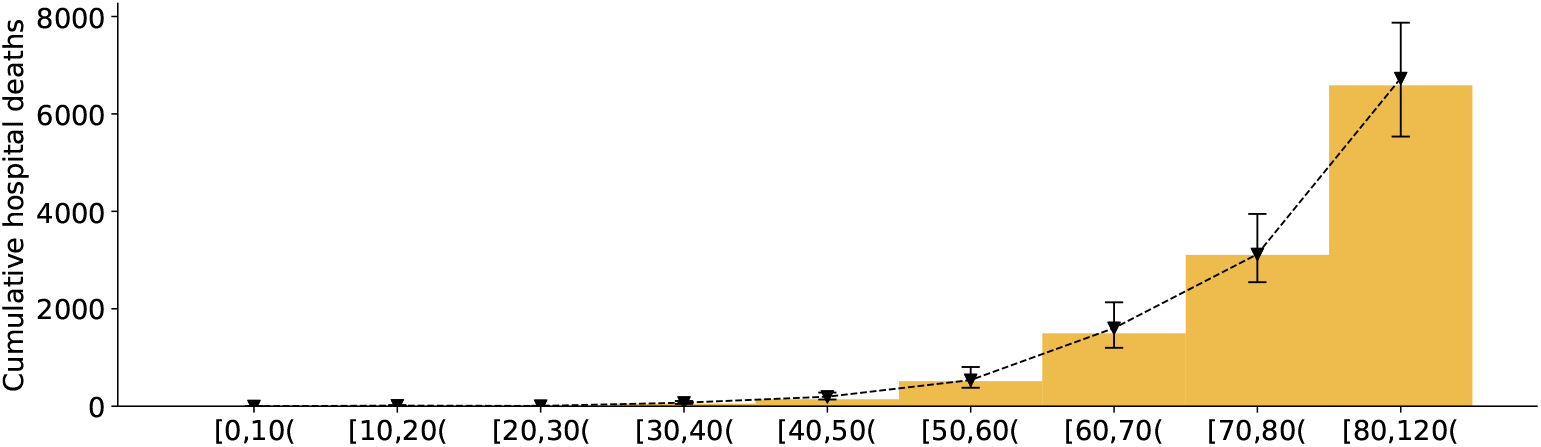
Cumulative deaths in Belgian hospitals per ten-year age strata. For the second Belgian 2020 COVID-19 wave, from September 1st, 2020 until February 1st, 2021. Yellow bars represent the data collected by Sciensano, inverted triangles represent the model prediction mean with 95 % confidence interval.

**Figure 13:**
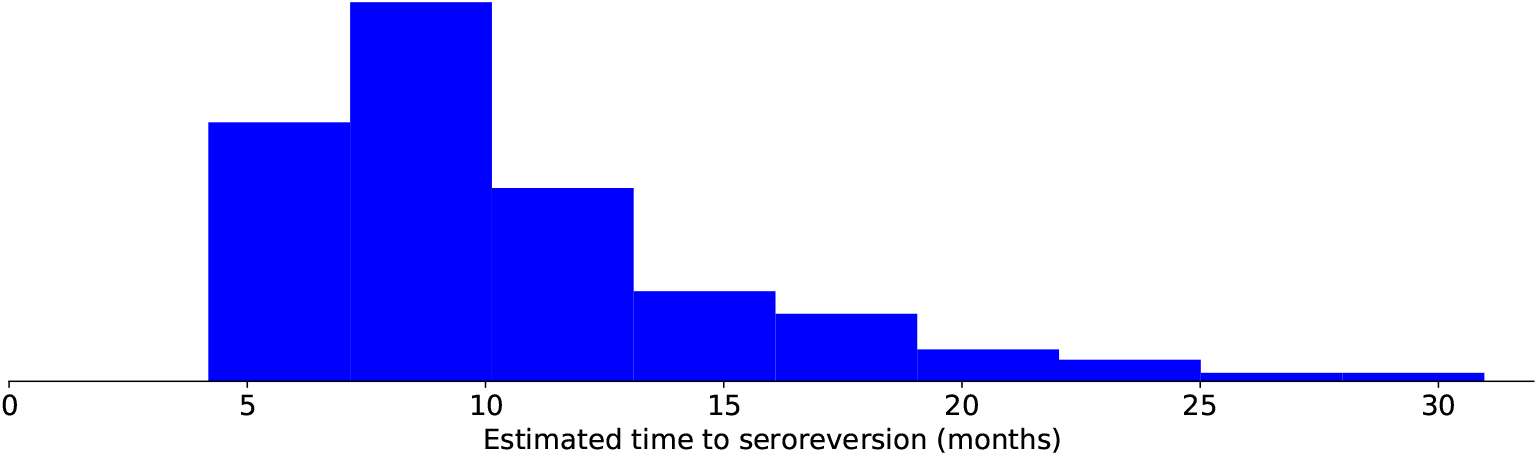
Estimated distribution of the time to seroreversion (1*/ζ*). The mean time to seroreversion is 9.2 months (IQR: 7.2 months - 12.1 months).

**Figure 14:**
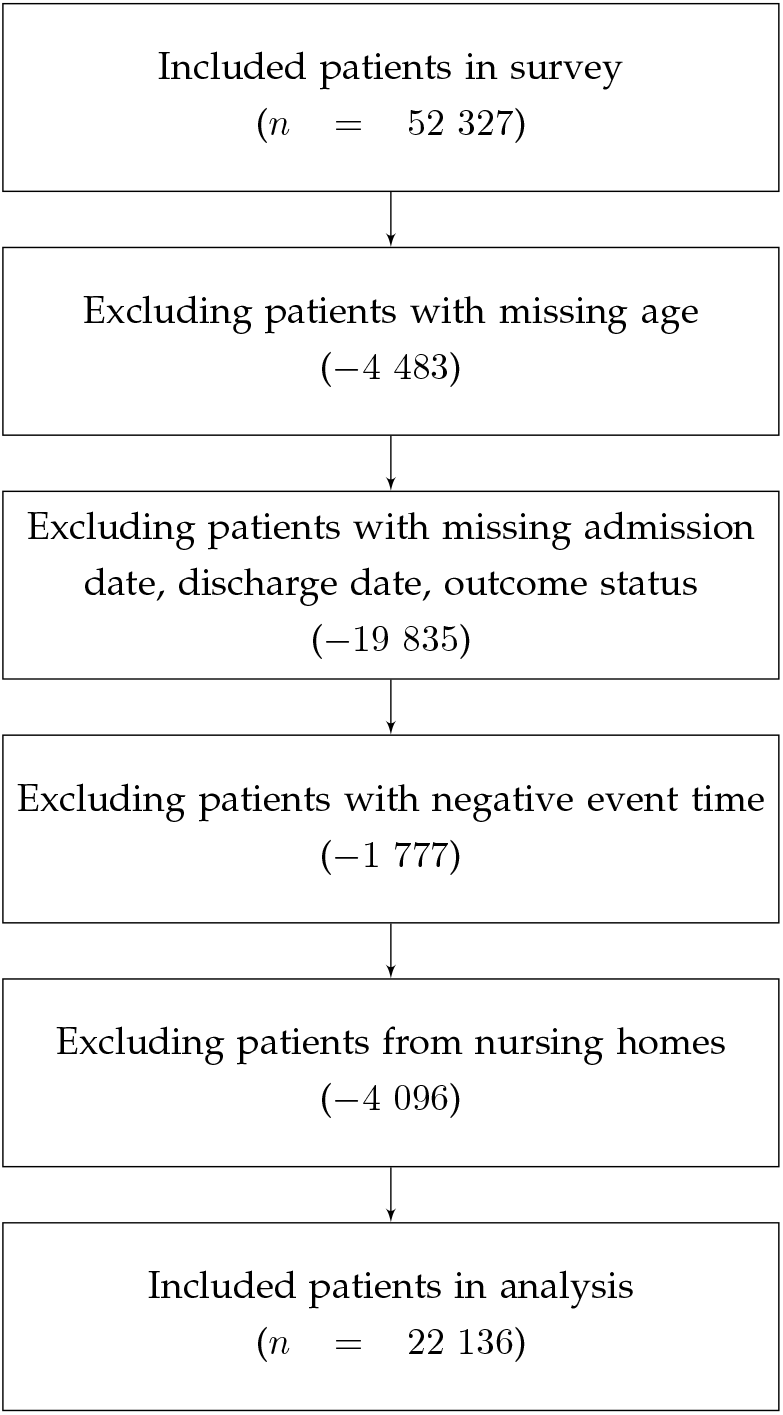
Flow diagram illustrating the number of patient data excluded from the survey and the reason thereof.

**Figure 15:**
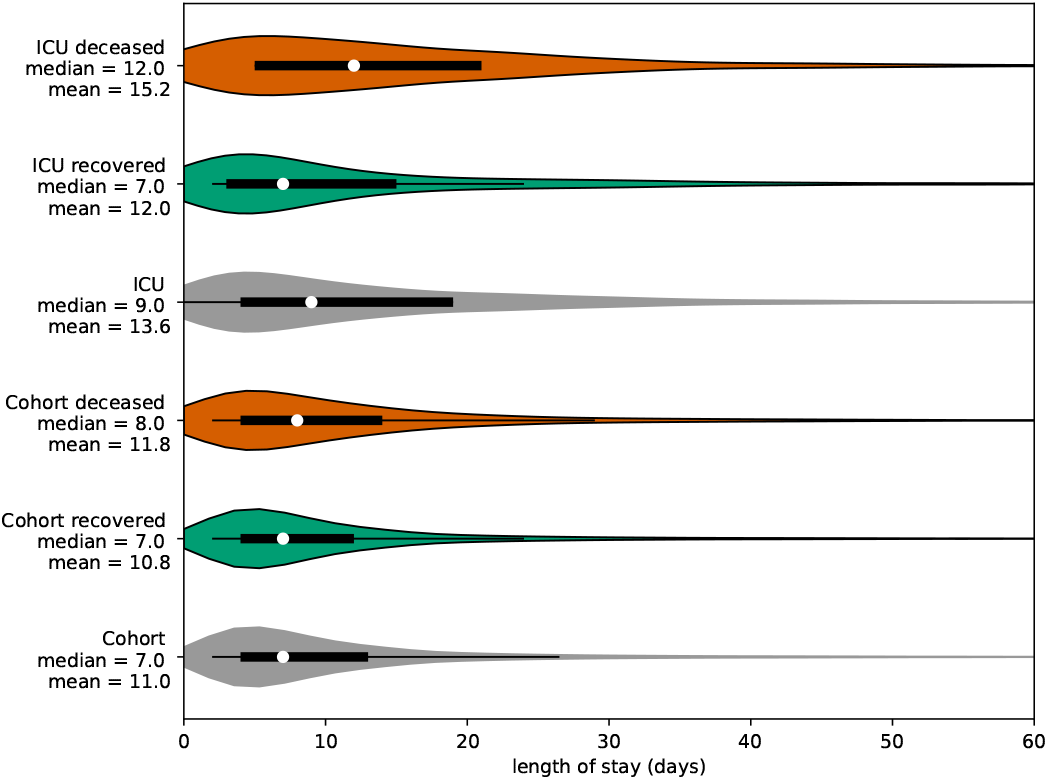
Observations of the length of a hospital stay for patients in cohort and ICU wards. Overall (gray), if recovered (green), if deceased (red). Residence times in cohort are shorter than residence times in ICU. In both wards, recovered patients have longer stays than deceased patients.

**Figure 16:**
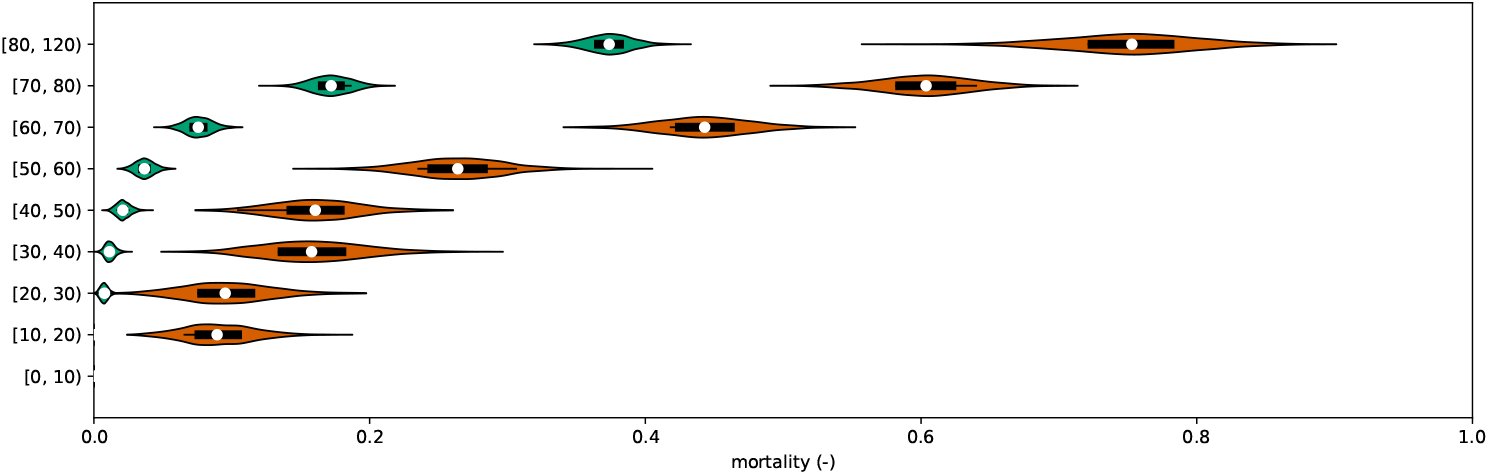
Mortality in cohort (***m***_C_, green) and mortality in ICU (***m***_ICU_, red) per ten-year age strata. Obtained by bootstrap resampling of the Belgian COVID-19 clincial surveillance on hospitalizations by Van Goethem et al. [40]. Mortality in both wards increases with patient age, mortality in ICU is higher than mortality in cohort.

**Figure 17:**
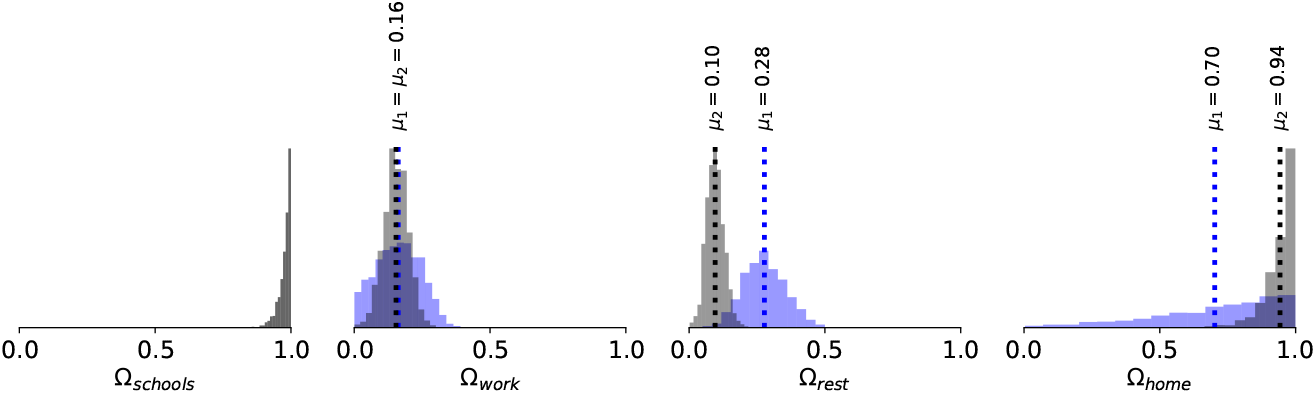
Inferred effectivity parameters at home (Ω_home_), in the workplace (Ω_work_), in schools (Ω_schools_) and for the sum of leisure activities, other activities and public transport (Ω_rest_), for the first COVID-19 wave (blue) and for the second COVID-19 wave (black). The effectivity of contacts in schools could not be deduced during the first COVID-19 wave because schools remained practically closed until July 1st, 2020. However, a high effectivity of contacts in schools could be deduced during the second COVID-19 wave. The effectivity of work contacts was roughly the same during both 2020 COVID-19 waves. The effectivity of leisure contacts was estimated to be lower during the second COVID-19 wave, however, leisure policies were not varied (yet) during the second COVID-19 wave, so the estimate must be taken with a grain of salt. Home contacts were deemed more effective by the model during the second COVID-19 wave.

### 4.2 Model calibration

We obtained an average basic reproduction number of *R*_0_ = 4.16 (IQR: 3.90 - 4.39) for the first 2020 COVID-19 wave and of *R*_0_ = 3.69 (IQR: 3.64 - 3.75) for the second 2020 COVID-19 wave, which is in line with the global consensus range of *R*_0_ = [2, 4]. The estimate for the second COVID-19 wave is slightly lower, and this is most likely because this estimate implicitly includes the effects of preventive measures and mentality changes that were gradually adopted during the first 2020 COVID-19 wave. The compliance to social measures was similar between both 2020 COVID-19 waves, little lag was observed (0.22 vs. 0.39 days) and the time to reach full compliance was of the same magnitude (9.17 vs 6.94 days). Thus, compliance to lockdown restrictions can be modeled using a ramp function without lag, eliminating one of the model’s parameters, namely *τ* (Equation 18). The seroreversion rate was estimated using two serological datasets. The data by Herzog et al. [39] consists of residual blood samples sent to laboratories, while the dataset of Sciensano consists of blood samples from Red Cross blood donors. The dataset of Herzog et al. [39] is likely biased towards sick individuals, while the dataset of Sciensano is biased towards healthy individuals. In the calibration procedure, both datasets were given equal weights to incorporate a *truth in the middle* heuristic. We estimated the average time to seroreversion as 9.2 months (IQR: 7.2 - 12.1 months), although the estimated distribution (Figure 13) has a long tail, it is very likely that antibody immunity gradually wanes. The estimate is consistent with the finding that 50 % of antibodies are most likely lost one year after the infection [24]. Using the same dataset, Abrams and colleagues [11] have estimated the rate of antibody waning at 8 months using their SARS-CoV-2 model (informal communication). It should be noted that the incorporation of antibody waning completely ignores the effects of cellular immunity and that more research on the exact kinetics of the immune response is necessary. In spite, it is best to include waning immunity in SARS-CoV-2 models, especially when long time-horizons are considered in the simulations. In this study, a population average subclinical fraction of 57 % was used, which was higher than estimated in a systematic review by Buitrago-Garcia et al. [36] (31 %) and higher than the estimate for the Icelandic population of Gudbjartsson et al. [35] (43 %). We expect that a decrease in the subclinical fraction could be compensated by a decrease of the per-case hospitalization risk (***h***) to obtain the same fit to the hospitalization data. However, lowering the subclinical fraction would lead to a reduced fraction of seropositive individuals in the population and thus a mismatch between the simulated seroprevalence data and observed seroprevalence data. Because of the good agreement between the simulated and observed seroprevalence, the fraction of subclinical infections are most likely correctly represented in the model (Figures 9 and 10).

We calibrated the model’s effectivity parameters (Ω_home_, Ω_schools_, Ω_work_, Ω_rest_) on incrementally larger hospitalization datasets and found that the model provides accurate forecasts under the observed mobility changes, even when the posteriors still depend on the extent of the dataset. However, *correct*^2^*effectivity parameters could only be deduced a posteriori events. This is because information* on the effectiveness of contacts can only be obtained by observing the hospitalizations under changing policies. Examples are the effects of leisure and work relaxations during the first COVID-19 wave and the effect of schools re-opening during the second COVID-19 wave. From April 15th, 2020 onwards (Figure 5, panel b) the ever decreasing trend in the daily hospitalizations is nicely captured even with posteriors seemingly converging to distributions different than those of the maximal dataset (panel f). Still, on May 1st 2020 (panel c), the model could have been used to accurately inform policymakers on the effects of lifting work and leisure restrictions just four days later. As soon as restrictions are lifted, the posteriors quicly converge to their final distributions. A similar observation is made with regard to the schools effectivity parameter. From November 7th, 2020 onwards (Figure 6, panel a) the effect of schools re-opening is captured in the model uncertainty, in spite of deviant posterior distributions. From Decenber 18th, 2020 onwards (panel c) the effect of schools re-opening is captured both in the model predictions and the effectivity parameters. Because accurate posteriors can only be inferred a posteriori, the modeler must asses if policy changes have been sufficient to deduce meaningfull effectivity posteriors. This is important when performing scenario analysis, as incomplete knowledge of the effectivity posterior can significantly alter the results.

Scaling pre-pandemic contact matrices with public mobility data has proven to be a rapidly deployable and cheap alternative to the use of survey-based contact studies under lockdown measures, such as the one of Coletti et al. [17] for Belgium. The social contact model is well-fit for the acute stages of the pandemic when these contact data are still being gathered. However, as the pandemic progresses, the survey-based contact studies are the preferred choice as the use of public mobility data is more coarse-grained. Because the GCMRs are not available for different age groups, they do not allow us to accurately capture how individuals of different ages have altered their behavior under lockdown measures. For example, the contact study by Coletti et al. [17] shows that younger individuals tend to increase their contacts sooner than older individuals after the release of lockdown measures. These differential effects are still captured in our social contact model, albeit less accurate than the survey-based contact model, by the multiplication of the GCMRs with the pre-pandemic number of contacts. For example, the mobility reduction in workplaces is only applied to the matrix of work contacts, which only contains contacts for individuals between 20 and 60 years old. Further, because the GCMRs are collated smartphone data, one could expect the elderly population to be underrepresented due to lower smartphone usage. However, it is unlikely that this would drastically alter our study’s results because older individuals have fewer contacts than younger individuals and thus contribute less to overall SARS-CoV-2 spread.

### 4.3 Effects of non-pharamaceutical interventions

Finally, we would like to discuss the importance of schools in the SARS-CoV-2 pandemic. As previously mentioned in section 3.3, there seems to be a strong correlation between school re-opening, the rise of laboratory-confirmed cases amongst youths, the rise of the number of clusters in schools, and the emergence of plateaus in the daily hospitalizations (Figures 4 and 7). Our model incorporates this correlation as high effectivities of school contacts. An increase in the effective reproduction number, from *R*_*e*_ = 0.66 ± 0.04 to *R*_*e*_ = 1.09 ± 0.05, is observed when schools are re-opened. Several studies have found children to be less susceptible to a SARS-CoV-2 infection [3, 26, 51]. Because quantitative data was scarce at the time of writing, we incorporated no changes in susceptibility and infectiousness in children in this study. However, this will not alter the large impact schools seem to have on SARS-CoV-2 spread in our model. If the susceptibility and infectiousness in children is lowered, this will most likely be countered during the parameter inference, where we expect higher values for the effectivity of contacts of children in schools (Ω_schools_) to be inferred. Although the present evidence is circumstantial, and correlation does not imply causation, schools seem to play a critical role in SARS-CoV-2 spread. Thus, school closure seems an effective way of countering an epidemic SARS-CoV-2 trend.

## 5 Conclusions

We obtained an average basic reproduction number of *R*_0_ = 4.16 (IQR: 3.90 - 4.39) and *R*_0_ = 3.69 (IQR: 3.64 - 3.75) for both 2020 COVID-19 waves in Belgium. We found that SARS-CoV-2 strongly discriminates between individuals of different age groups, with youths and the working-aged population driving the pandemic, and the senior population needing hospital care. These results are in line with the established consensuses and highlight the model’s validity. Further, by propagating the hospitalization parameters computed using the clinical surveillance dataset, the model is able to accurately predict the number of daily hospitalizations, the total number of patients in Belgian hospitals, the total number of deaths in Belgian hospitals, and the seroprevalence in the Belgian population during both 2020 COVID-19 waves.

The combination of the deterministic epidemiological model, which incorporates a-priori knowledge on disease dynamics, and the social contact model whose infectivity parameters were inferred allow us to make the most out of the available pre-pandemic data and public mobility data. Our method is computationally cheap and does not require ad-hoc tweaking to obtain a good fit to the observed data. A disadvantage is that the effectivity parameter distributions only converge to their *correct* posterior distributions a posteriori policy changes. Still, even when using a very limited calibration dataset, the model is able to make accurate predictions of the future number of hospitalizations, highlighting the robustness of the calibration method.

As soon as schools were re-opened on November 16th, 2020, the number of confirmed cases amongst youths starts increasing. A significant lead relationship between the number of cases amongst youths and the working population, and youths and the senior population was found. Our model incorporates this correlation as high effectivities of school contacts. When schools were re-opened under lockdown policies, the model indicates the effective reproduction number increased from *R*_*e*_ = 0.66 ± 0.04 to *R*_*e*_ = 1.09 ± 0.05. Thus, school closure is an effective measure to counter an epidemic SARS-CoV-2 trend.

## Data Availability

All code and data is publically available on the BIOMATH GitHub repo (https://github.com/UGentBiomath/COVID19-Model/).

## 6 Future research

- The calibration procedure should be repeated using pandemic social contact matrices, which are currently being gathered for Belgium by Coletti et al. [17]. Further, the effects of integrating the contacts with their duration should be explored. A comparison between the different results can then be made.
- The effective reproduction number in the different places should be compared to data on SARS-CoV-2 clusters to further validate the model.
- It is expected that lockdown measures in Belgium will be lifted soon. The impact of releasing measures on the daily hospitalizations should be studied to find a link between the effectivity parameters and the mobility reductions.
- If schools are a major contributor to SARS-CoV-2 spread, administering a vaccine with high transmission-blocking potential to youths is expected to have a similar effect as schools closure. Due to their localized nature, vaccination for SARS-CoV-2 in schools is logistically easier than vaccinating the general population.

## Acknowledgements

This work is the result of a team effort. I want to thank Daniel Illana, Bram De Jaegher, Daan Van Hauwermeiren, Stijn Van Hoey and Joris Van den Bossche for their help in maintaining the GitHub repo, coding the visualizations and for teaching me the basics of object-oriented programming in Python. I would like to thank Mieke Descheppere, from the Ghent University hospital and Wim Verbeke, MD from AZ Delta Roeselare, for sharing their insights on hospital dynamics. We thank *VZW 100 km Dodentocht Kadee* for their financial support through the organisation of the 2020 100 km COVID-Challenge.

## Ethics statement

Permission for the clinical hospital surveillance was granted by the Committee on Medical Ethics of the Ghent University Hospital (BC-07507) and by the Belgian Federal Information Security Committee to the Belgian Collaborative Group on COVID-19 Hospital Surveillance. The need for informed consent was renounced, however, upon hospital discharge, patients were informed that their data would be curated by the Belgian Scientific Institute for Public Health (Sciensano) and used in the context of the COVID-19 public health crisis for policy-supporting research.

## CRediT author statement

**Tijs W. Alleman:** Conceptualization, Software, Methodology, Investigation, Data Curation, Writing - Original Draft. **Jenna Vergeynst:** Conceptualization, Software, Writing - Review. & Editing, Project administration. **Lander De Visscher:** Methodology. **Michiel Rollier:** Methodology, Writing - Review. **Elena Torfs:** Conceptualization, Funding acquisition. **Ingmar Nopens:** Conceptualization, Funding acquisition, Project administration, Writing - Review. & Editing **Jan Baetens:** Conceptualization, Funding acquisition, Project administration, Writing - Review & Editing. **Belgian Collaborative Group on COVID-19 Hospital Surveillance** Data collection, Data Curation.

## A Supplementary materials

### A.1 Overview of model assumptions and limitations

The following assumptions were made concerning the SEIQRD dynamics:

1. All individuals experience a brief presymptomatic, infectious period.
2. All individuals, including children, are equally susceptible to SARS-CoV-2 infection. It is unlikely that lower susceptibility in children would alter the dominant role of schools in SARS-CoV-2 transmission. During model calibration, a higher effectivity of the contacts in schools (Ω_schools_) could compensate for the lower susceptibility in children.
3. Asymptomatic and mild cases automatically lead to recovery and in no case to death.
4. Mildly infected and hospitalized individuals cannot infect susceptibles (= *quarantined*). A fraction of individuals experiencing influenza-like illness will not reduce their number of non-household contacts and will thus contribute to disease spread [52]. In our model, this behavior is not accounted for. The model cannot be used to model the effect of transmission to healthcare workers.
5. All deaths come from hospitals, meaning no patients died at home [53].
6. The modeled population is the general population of Belgium and does not explicitly take nursing homes into account. The model is unfit to make predictions on nursing home deaths.
7. Waning of antibody immunity is incorporated in the model as individuals transitioning from the recovered (R) population pool to the susceptible (S) population pool. The incorporation of antibody waning ignores the effects of cellular immunity (through T-and B-cells). More research on the exact kinetics of the immune response is necessary to finetune to the model.

The following assumptions to the hospital dynamics were made:

1. Upon arrival in the hospital, all patients immediately transfer to a cohort ward or an ICU. In real life, a patient may first spend some time in a cohort ward before going to an ICU and this is not accounted for.
2. Residence times in cohort and in ICU differ depending on the outcome of the infection (recovered or deceased).
3. All recovered ICU patients spend some additional time in cohort (recovery and observation stay).
4. Patients in nursing homes were excluded from the analysis of the clinical surveillance dataset. The model can make predictions on hospital deaths in individuals coming from the general population.
5. During the analysis of the hospital surveillance data, the data analysis was not split into several time intervals and hence the temporal changes in hospital residence times and mortalities were neglected. In spite, Faes et al. [41] have reported that the median residence time decreased after the first 2020 COVID-19 wave.

The following assumptions were made in the social contact model:

1. Prepandemic contact matrices by Willem et al. [14] are scaled with mobility reductions extracted from the GCMRs and an effectivity parameter inferred from hospitalization data using a *Markov-Chain Monte-Carlo* method to mimic pandemic social behavior.
2. The GCMRs are not age-stratified and do not correct for a potential underrepresentation of older individuals in the data collection. The GCMRs are a more coarse-grained approach as compared to social-epidemiological contact studies that estimate mixing patterns under lockdown measures [17]. However, setting up a survey-based contact study is a resource and time-intensive endeavor. The advantage of using the GCMRs in our social contact model is their rapid and public availability, making their use appropriate during the early stages of a pandemic when more accurate survey-based contact studies are being set up.
3. The effectivity of the contacts (Ω_*x*_) are bound between zero and one. This implies that if work mobility is reduced to 40 % of its pre-pandemic value, the work contacts can account for no more than 40 % of its pre-pandemic value.
4. There is no link between the effectivity parameters and the mobility reduction. However, when relaxing measures, an increase in mobility will likely be accompanied by an increase in the effectiveness of school contacts. This is due to mentality changes upon relaxation, as measures will gradually be ignored more.

### A.2 Overview of model parameters

### A.3 Key events

The first lockdown, which started on March 15th, 2020, and lasted until May 4th, 2020 involved the closure of schools, bars, clubs, restaurants, all non-essential shops, and closure of the border to non-essential travel (Table 2). The GCMRs show a 56 % reduction in work-place mobility (Figure 2 and Table 2). Based on surveys from the Belgian National Bank, 28.6 % of all employees were able to work from home, 29.9 % remained in the workplace and 4.4 % worked both from home and in the workplace. 32.4 % were temporary unemployed and 4.8 % were absent [55]. Public transport mobility decreased by 65 %, leisure mobility decreased by 72 %, and grocery & pharmacy mobility was reduced by 26 %. From March 15th, 2020 until May 4th, 2020, mobility remained practically constant at the aforementioned reductions. On May 4th, 2020 the lockdown was gradually lifted by re-opening all non-essential shops and lifting telework restrictions. The effect can be seen in the *Google Community Mobility Reports* (Figure 2), by the end of April, workplace and retail & recreation mobility gradually start increasing. By July 1st, 2020, almost all social measures had been lifted. During the first lockdown, schools remained fully closed until May 18th, 2020, and were only re-opened to a very limited extent before the end of the school year on July 1st, 2020. For this reason, schools are assumed to remain closed during the first COVID-19 wave. During July, there were few social restrictions, and this resulted in new, localized infection clusters. During most of August 2020, a lockdown with a curfew was imposed in Belgium’s Antwerp province. We do not attempt to model the hospitalizations during July and August 2020, as modeling localized infection clusters with a nation-level epidemiological model can only be accomplished by severe ad-hoc tweaks in the social contact model. A spatial model extension was developed to better account for such localized phenomena.

During the second lockdown from October 19th, 2020 until the present day (26/02/2021), workplace mobility has been reduced by approximately 25 %. During Autumn break and Christmas holidays, workplace mobility further declined to approximately 45 %. Public transport mobility decreased by 30 % and by 50 % during holidays, leisure mobility decreased by 40-50 % and grocery & pharmacy mobility have decreased by approximately 5-10 %. Primary and secondary schools were closed between October 19th, 2020, and re-opened on November 16th, 2020. Further, schools have been closed during the Christmas holidays from December 18th, 2020 until January 4th, 2021, and were closed during spring break from February 15th, 2021 until February 21th, 2021. Universities have remained fully closed since October 19th, 2020.

During both lockdowns, increases in the categories *residential* and *parks* were observed (Figure 2). These are indicative of decreased mobility, as these suggest increased activity around the home environment. The other four categories are more indicative of general mobility as they are related to activity around workplaces, retail outlets and use of public transportation [43]. Thus, although the mobility figures indicate people spent more time at home, this does not mean people have more contacts at home (especially under stay-at-home orders). Amplifying the fraction of household contacts under lockdown measures would increase intergenerational mixing of the population under lockdown, which is unrealistic and will lead to overestimations of the hospitalizations. The inability to accurately capture the disease spread in home *bubbles* under lockdown measures is an inherent downside of compartmental epidemiological models. We have thus not scaled the home interaction matrix (***N***_c,home_) with the residential mobility from the GCMRs.

### A.4 Basic reproduction number

Since the system of differential equations (Eq. 1 - Eq. 12), is autonomous, the eigenvalues of the Jacobian matrix evaluated at its hyperbolic equilibrium point can be used to determine the nature of that equilibrium [64]. The basic reproduction number (*R*_0_) is computed as the spectral radius of the Jacobian matrix at the disease-free equilibrium [27]. Our model has seven infected states: *E, I*_presy_, *I*_asy_, *Q*_mild_, *Q*_cohort_, *Q*_ICU_ and *Q*_ICU, rec_ (Figure 1). At the disease-free equillibrium, the whole population is susceptible to the infectious disease, *S*_*i*_ *T*_*i*_,

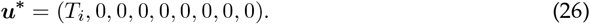

The Jacobian ***J*** is defined as,

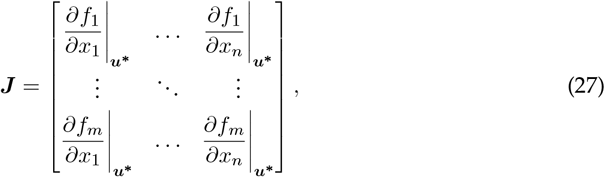

where *n* and *m* are equal to the number of infected compartments. Next, the Jacobian is decomposed in the following form,

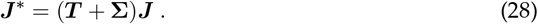

The matrix ***T*** contains all terms that lead to *transmissions* of SARS-CoV-2, while **Σ** contains all terms that lead to *transitions*. For our model,

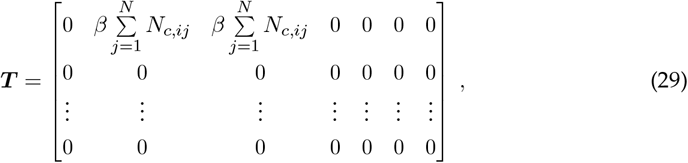

where an entry *T*_*i,j*_ is the rate at which individuals in infected state *j* gives rise to individuals in infected state *i*. And,

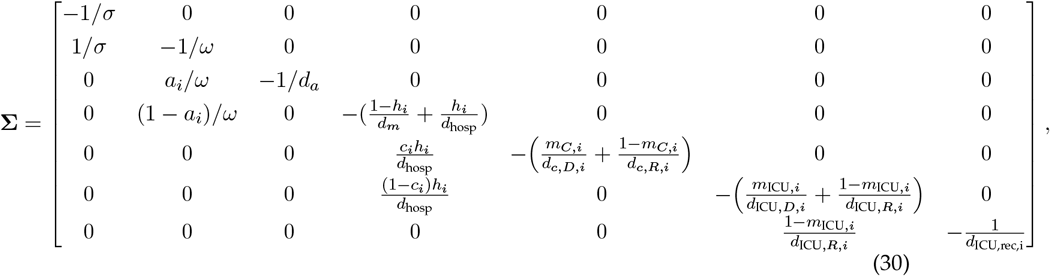

where an element 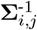 is the expected time that an individual who presently has state *j* will spend in state *i* during its entire epidemiological *life*. The next generation matrix (NGM) is then calculated as,

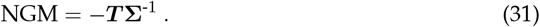

The basic reproduction number *R*_0_ is defined as the spectral radius^3^ ***ρ*** of this matrix [27],

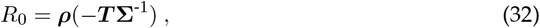

which becomes for our model,

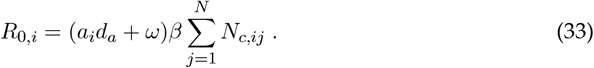

A linear relationship between the reproduction number and the chance of infection upon contact (*β*), the number of contacts (***N***_***c***_) and the sum of the durations of infectiousness for those compartments able to infect susceptibles makes sense.

### A.5 Time-lagged cross correlation

We extracted the number of laboratory confirmed cases in youths [0, 20[, the working population [20, 60[ and the senior population [60, ∞[ from the *Belgian Scientific Institute of Public Health* (https://epistat.sciensano.be/Data) from November 2nd, 2020 to February 1st 2020. We then normalized the timeseries with the number of cases on November 21st, 2020 and visualized the result in Figure 4. Using the Python module *pandas*, the dataseries were shifted with *k* days and the cross correlation was computed. The procedure was performed for *k* ∈ [−15, 5] days, the resulting *cross correlation function* is shown in Figure 8 and the results of the analysis are summarized in Table 3. Next, we constructed a statiscal test to check if the covariance between two series *x* and *y*, shifted with the number of days resulting in the maximum covariance, *k*_*max*_, varied significantly from zero. Thus, the null hypothesis is,

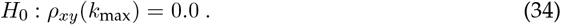

If the cross correlation of lag *k*_max_ is zero, then, for a fairly large timeseries consisting of *n* datapoints, the covariance *ρ*_*xy*_(*k*_max_) will be approximately normally distributed, with mean zero and standard deviation 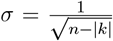. Since approximately 95% of a normal population is within 2 standard deviations of the mean, a test will reject the hypothesis that the cross correlation of lag *k* equals zero when,

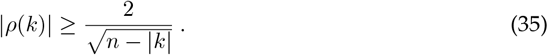

The null hypothesis was rejected for all timeseries.

### A.6 Supplementary data and figures

## Footnotes

1 Date of school reopening 2021-11-16 plus one five-day incubation period.

2 Assuming the inferred posterior distributions of the maximal dataset are correct.

3 Largest absolute eigenvalue.

